# SARS-CoV-2 correlates of protection from infection against variants of concern

**DOI:** 10.1101/2024.05.28.24308095

**Authors:** Kaiyuan Sun, Jinal N. Bhiman, Stefano Tempia, Jackie Kleynhans, Vimbai Sharon Madzorera, Qiniso Mkhize, Haajira Kaldine, Meredith L McMorrow, Nicole Wolter, Jocelyn Moyes, Maimuna Carrim, Neil A Martinson, Kathleen Kahn, Limakatso Lebina, Jacques D. du Toit, Thulisa Mkhencele, Anne von Gottberg, Cécile Viboud, Penny L. Moore, Cheryl Cohen, PHIRST-C group

## Abstract

Serum neutralizing antibodies (nAbs) induced by vaccination have been linked to protection against symptomatic COVID-19 and severe disease. However, much less is known about the efficacy of nAbs in preventing the acquisition of infection, especially in the context of natural immunity and against SARS-CoV-2 immune-escape variants. In this study, we conducted mediation analysis to assess serum nAbs induced by prior SARS-CoV-2 infections as potential correlates of protection (CoPs) against Delta and Omicron BA.1/2 wave infections, in rural and urban household cohorts in South Africa. We find that, in the Delta wave, anti-D614G nAbs mediate 37% (95%CI 34% – 40%) of the total protection against infection conferred by prior exposure to SARS-CoV-2, and that protection decreases with waning immunity. In contrast, anti-Omicron BA.1 nAbs mediate 11% (95%CI 9 – 12%) of the total protection against Omicron BA.1/2 wave infections, due to Omicron’s neutralization escape. These findings underscore that CoPs mediated through nAbs are variant-specific, and that boosting of nAbs against circulating variants might restore or confer immune protection lost due to nAb waning and/or immune escape. However, the majority of immune protection against SARS-CoV-2 conferred by natural infection cannot be fully explained by serum nAbs alone. Measuring these and other immune markers including T-cell responses, both in the serum and in other compartments such as the nasal mucosa, may be required to comprehensively understand and predict immune protection against SARS-CoV-2.

## Main text

The acute phase of the COVID-19 pandemic has waned with the development of SARS-CoV-2 population immunity in most individuals through repeated episodes of vaccination, infection, or both ^1,2^. Owing to the unprecedented speed of SARS-CoV-2 vaccine development and distribution ^3^, considerable numbers of people were primed by vaccination, averting substantial morbidity and mortality ^4^. However, due to immune evasive variants, vaccine hesitancy, and lack of global equity in vaccine access ^5–7^, a substantial proportion of the world’s population acquired SARS-CoV-2 immunity through natural infections, especially in low- and middle-income countries ^8,9^. Immune markers that reliably predict protection from infection or symptomatic disease are known as “correlates of protection” (CoP). The post-pandemic era is marked by rapid antigenic drift of Omicron subvariants leading to continued immune evasion ^10–13^. Given this complex evolutionary landscape, it remains important to identify CoPs induced by natural infections and/or vaccinations against SARS-CoV-2 variants to monitor population susceptibility, anticipate future waves, optimize rollout of existing vaccines, and facilitate design and approval of next generation vaccines ^14^. There has been significant progress in defining serum neutralizing or binding antibodies to the spike protein as CoPs for COVID-19 vaccines, although most of the data are derived from early randomized controlled trials focused on peak immune response shortly after vaccination and measured against symptomatic disease caused by the ancestral strain, with updated data on variants ^15–24^. In comparison, little is known about serum CoPs for infection-induced immunity and protection against acquisition of subclinical infections.

CoPs may differ for immunity induced by infection vs. vaccination: SARS-CoV-2 infections tend to induce more robust mucosal immunity despite lower serum antibody responses than intramuscularly delivered mRNA vaccines, as shown in a mouse model ^25^, and mucosal immunity may play a more important role in reducing risk of infection and transmission than systemic immunity ^26,27^. Moreover, CoPs need to be interpreted in the context of viral evolution: in the pre-Omicron era, SAR-CoV-2 variants of concerns emerged independently from one another, with the Alpha, Beta, Gamma, Delta, and Omicron variants exhibiting distinct phenotypic characteristics. The Omicron variant stands out due to substantial genetic divergence from earlier strains and significant immune evasion capabilities against antibody neutralization ^28^. Equivalent antibody titers may not provide equivalent levels of protection against ancestral strains compared to more transmissible and immune-evasive variants like Omicron, and CoPs may therefore be variant-dependent. Furthermore, serum antibody titers against SARS-CoV-2 also wane with time.

The challenge of defining CoP for infection induced immunity partially stems from the difficulty of tracking immune exposures to SARS-CoV-2 infections, given that a significant proportion of infections are asymptomatic or subclinical and cannot be fully captured by traditional symptom-based surveillance protocols. The SARS-CoV-2, influenza, and respiratory syncytial virus community burden, transmission dynamics, and viral interaction in South Africa (PHIRST-C) cohorts in South Africa ^29,30^ overcame this challenge by implementing a rigorous sampling strategy, including collection of nasal swabs twice-weekly during a period of intense follow up, along with a total of 10 sequential blood draws spanning the D614G, Beta, Delta, and Omicron BA.1/2 waves. This high-intensity sampling scheme allowed us to reconstruct the cohort participants’ SARS-CoV-2 infection histories with high fidelity, and to monitor infection-induced antibody responses over time ^30^. Blood samples collected immediately prior to Delta and Omicron waves offered a unique opportunity to investigate serum immune marker levels in close proximity to the next SARS-CoV-2 exposure. Furthermore, vaccine-derived immunity remained low at the onset of the Omicron BA.1/2 wave, with less than 25% of the population fully immunized with Ad26.COV2.S (Janssen) and/or BNT162b2 (Pfizer BioNTech) vaccines ^31^. In this study, we leveraged the PHIRST-C cohorts’ unique serological and epidemiological data to perform mediation analysis and assess neutralizing antibody (nAb) titers induced by prior infection as CoPs against variants of concerns. Specifically, we evaluated the role of anti-D614G and anti-Omicron BA.1 nAbs against the Delta and Omicron BA.1/2 variants.

## Results

### Cohort description and antibody titer measurements

We analyzed data from the multi-year PHIRST-C cohort study, covering the first four waves of SARS-CoV-2 infections including the Delta and Omicron BA.1/2 waves ^29,30^. The study included a rural and an urban site in two provinces of South Africa. Households with more than two members and where at least 75% of members consented to participate were eligible. A total of 1200 individuals from 222 randomly selected and eligible households among the two study sites were longitudinally followed from June 2020 through April 2022. The study was characterized by intense nasopharyngeal swab and serum sample collection from the peak of the SARS-CoV-2 D614G (1^st^) wave to after the peak of the Delta (3^rd^) wave. After this initial follow-up period, nasopharyngeal swab sample collection stopped but serum samples continued with blood drawn immediately following the Omicron BA.1/2 (4^th^) wave. The timing of the serum sample collection is visualized in Fig. 1. We previously reconstructed the detailed SARS-CoV-2 infection history of each individual in the cohort up to the Omicron BA1/2 wave and demonstrated that immunity conferred by prior infection reduced the risk of reinfection ^30,32^. In this study, we extended this work to investigate how infection-induced neutralizing antibody (nAb) titers correlate with protection against SARS-CoV-2 reinfection with the Delta or Omicron BA.1/2 variants.

**Fig. 1:**
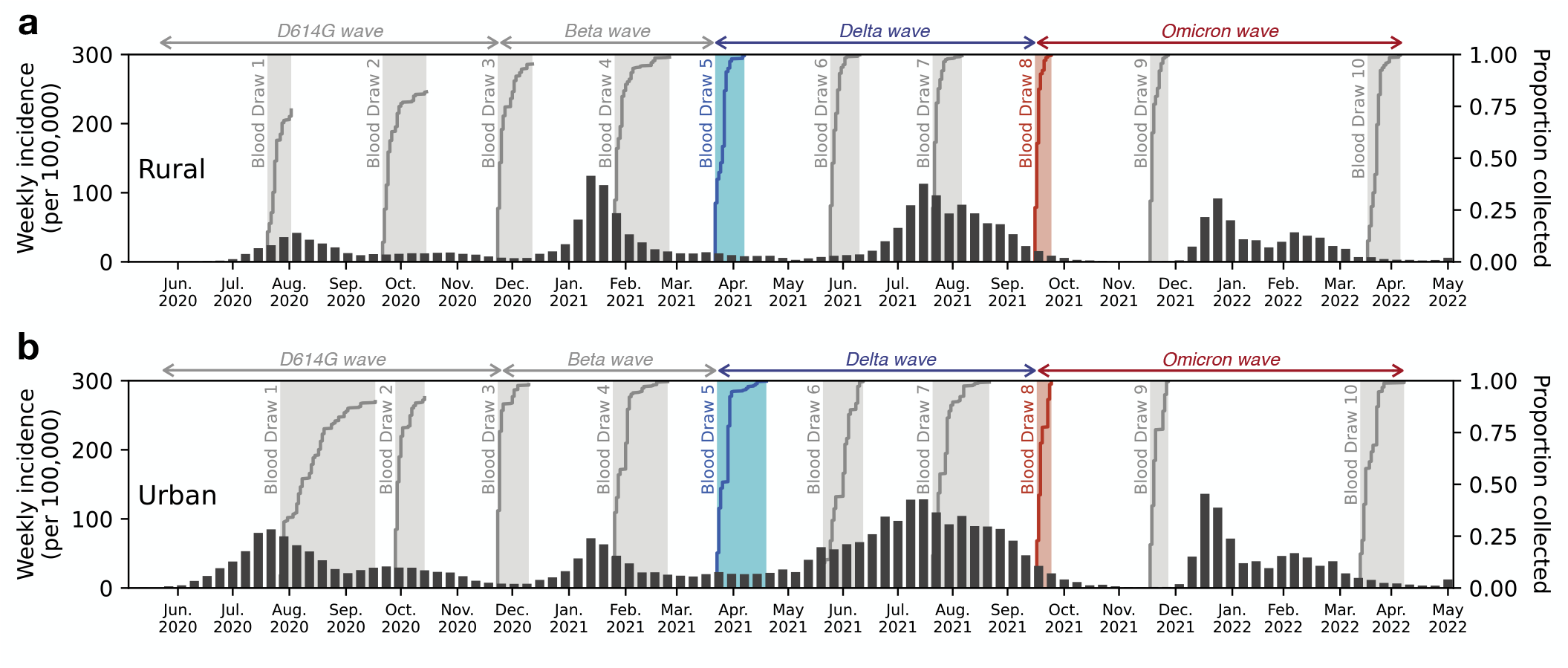
Timing of cohort sample collections with respect to SARS-CoV-2 variants’ circulations in the two study sites. **a**, Timing of the blood draws with respect to the SARS-CoV-2 epidemic waves in the rural site (Agincourt) of the PHIRST-C cohort. Bar plot represents the weekly incidence (per 100,000 population) of SARS-CoV-2 cases from routine surveillance data collected in Ehlanzeni District, Mpumalanga Province (where rural participants reside). The shaded areas represent the timing of the serum sample collections for the 10 blood draws. Each curve within the shaded area indicates the cumulative proportion of participants’ serum samples collected over time. The Delta wave subgroup analysis focuses on nAb titers among serum samples collected during blood draw 5 (blue shade); the Omicron BA.1 wave analysis focuses on nAb titers among serum samples collected during blood draw 8 (red shade). **b**, Same as (A), but for the urban site (Klerksdorp). The routine surveillance data (bar plot) were collected from Dr. Kenneth Kaunda District, North West Province (where urban participants reside).

For the Delta wave, we focused on a subgroup of 797 participants from 196 households (Delta wave subgroup, Table 1, Extended Data Fig. 1) who remained SARS-CoV-2 naïve or had a single prior SARS-CoV-2 infection before the Delta wave (hence, removing vaccinated and repeatedly infected individuals from the analysis; see Fig. 1 for the timing of the Delta wave). We define prior infection as positivity on the Roche Elecsys anti-nucleocapsid assay (an assay was optimized to detect prior infection ^33^), and/or rRT-PCR-positivity, at or before blood draw 5 (refer to BD5 hereafter). SARS-CoV-2 infections during the Delta wave were inferred based on the anti-nucleocapsid antibody level of two pre- and one post-Delta wave serum samples, as previously described ^30^. We focused on households with no more than six infected household members during this wave, due to computational constraints of the transmission model (see Methods for details). Among the 797 subgroup participants, 34% (273/797) were infected during the Delta wave, with attack rates of 42% (229/544) and 17% (44/253) for naïve and previously infected participants, respectively.

**Table 1:**
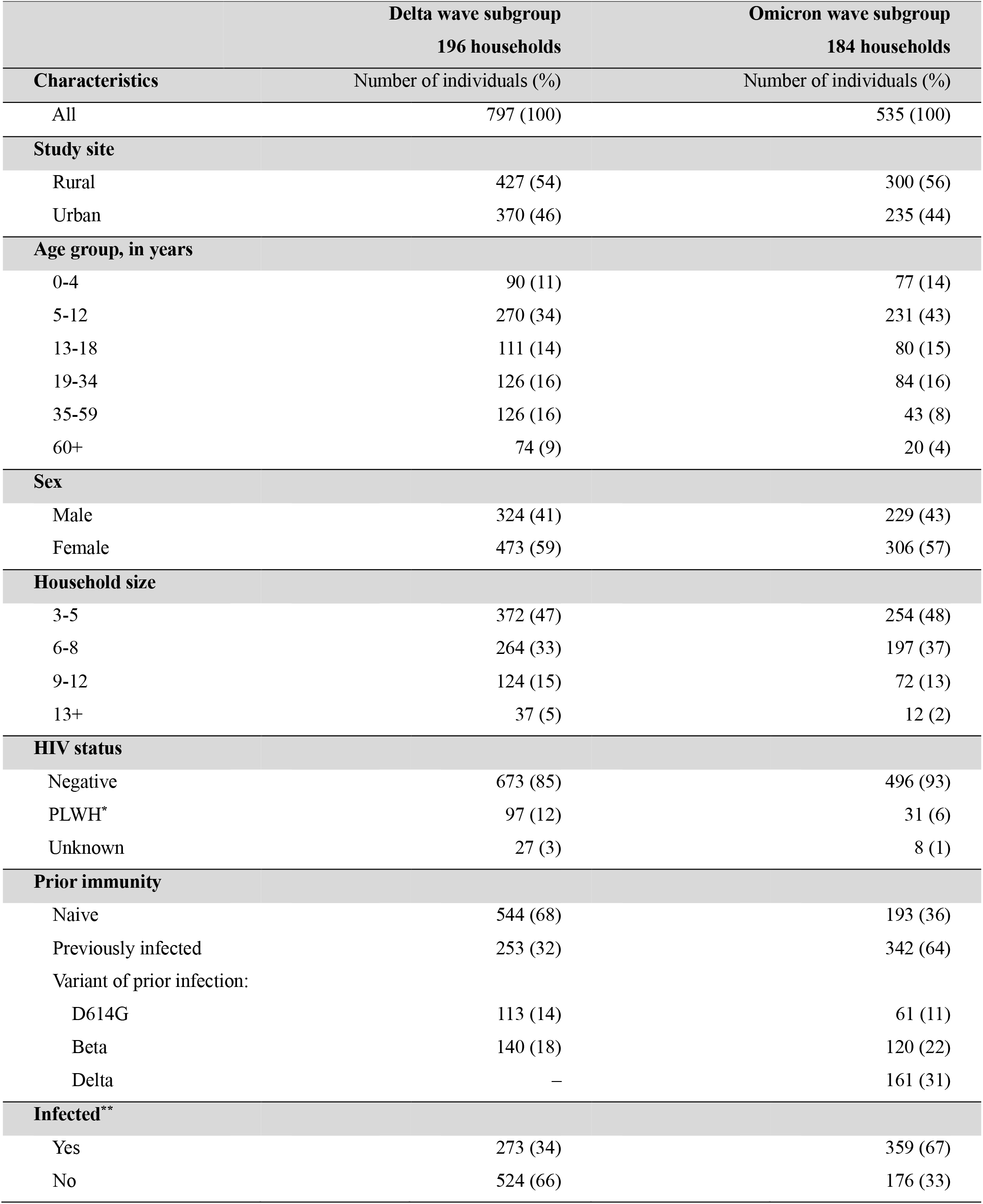
Characteristics of the PHIRST-C cohort’s Delta wave subgroup and Omicron wave subgroup populations, respectively. ^**\***^PLWH: people living with HIV. ^**^Here it indicates if a participant of the Delta/Omicron wave subgroup was infected (either primary or repeat infection) during the Delta/Omicron BA.1 wave.

To identify CoPs against the Delta variant, for the 253 participants who had been previously infected, we measured their anti-D614G nAb titers (measured as the inhibitory dilution at which 50% neutralization is attained, referred to as ID_50_ hereafter), using the blood draw immediately preceding the Delta wave (BD5). To evaluate the potential impact of antibody waning, we also measured the peak nAb level for each participant (defined as the highest anti-D614G nAb titer among the first 5 blood draws). We then calculated the degree to which nAbs had waned from peak level to that at BD5 by calculating the difference between peak nAb titer and nAb titer at BD5 (denoted as Δ*nAb*^*W*^ hereafter). If the peak response was already below the nAb assay detection threshold (which is set at 20), then Δ*nAb*^*W*^ was also assigned to be below the threshold, since further titer drop was not detectable. Notably, 28% (32/113) and 58% (81/140) of individuals previously infected with D614G and Beta exhibited anti-D614G nAb titers below the detection threshold at BD5, respectively (Extended Data Table 1). The proportion below the detection threshold was higher for individuals previously infected with the Beta variant than the D614G variant, given the Beta variant has eight amino acid difference in the spike, resulting in an antigenically distinct receptor binding domain compared to the D614G variant used in the neutralization assay. However, more than 90% of individuals remained positive on the Roche Elecsys anti-nucleocapsid assay for both prior D614G and Beta exposed individuals ^33^, despite low nAb titer level (Extended Data Table 1).

Fig. 2a shows the Delta wave participants anti-D614G nAb titers at peak and at BD5. The ID_50_ geometric mean titer (GMT) was 125 (95% CI 97 – 161) at peak and waned to 85 (95% CI 69 – 104) at BD5, representing an average 1.47 (95% CI 1.32 – 1.67) fold reduction due to waning. The anti-D614G nAb titers (in log scale) at peak and at BD 5 were highly correlated (Pearson correlation coefficient 0.89, p<0.0001). Comparing the nAb titers between individuals who were infected during the Delta wave vs. those who were not infected, we found that the GMTs of infected individuals was significantly lower than that of uninfected individuals for both anti-D614G nAb at peak level and at BD5 (Fig. 2b-c). In contrast, we did not find a significant difference in the degree of antibody loss due to waning (Δ*nAb*^*W*^) between infected and uninfected individuals (Fig. 2d).

**Fig. 2:**
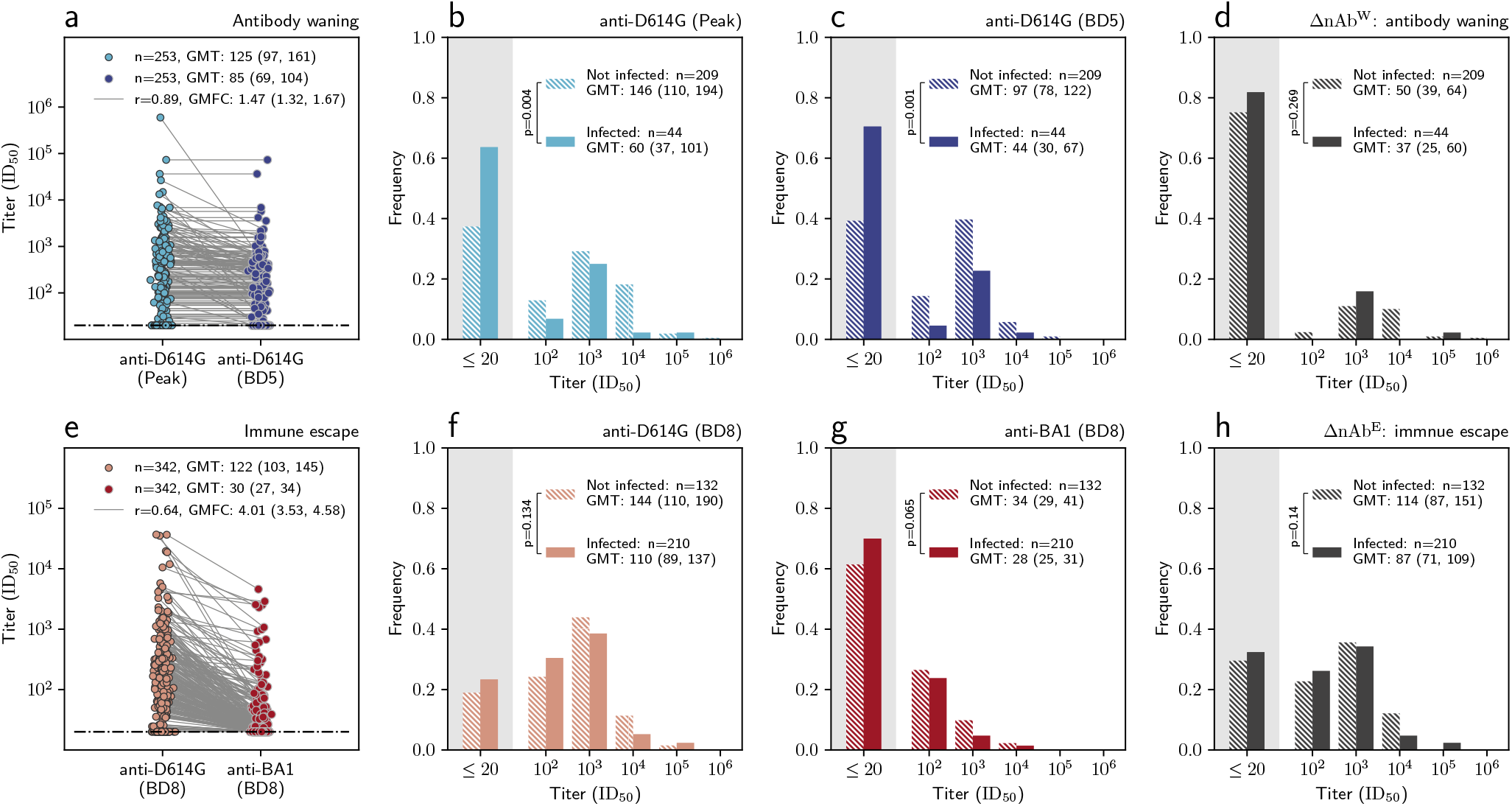
Anti-D614G and anti-BA.1 nAb titers for the Delta wave and the Omicron wave analysis. **a**, for Delta wave subgroup, the distribution of the peak anti-D614G nAb titer up to BD5 (light blue dots) and the anti-D614G nAb titer at BD5 (dark blue dots), among individuals who had one prior SARS-CoV-2 infection before blood draw 5. Each dot represents one individual, with two measurements of the same individual connected through a gray line. GMFC: geometric fold change from peak anti-D614G titer to that at BD5; GMT: geometric mean titer; r: Pearson correlation coefficient. **b**, for Delta wave subgroup, the distribution of the peak anti-D614G nAb titer up to BD5, stratified by individuals who were infected during the Delta wave (solid bar) vs those who were not infected (dashed bar). Independent samples t-test (two-sided) is used to determine the statistical significance (p-value reported on the legend) of difference between the GMT of the two groups. **c**, same as **b** but for anti-D614G nAb titers at BD5. **d**, same as **b** but for Δ*nAb*^*W*^ (defined as the difference between anti-D614G titers at peak and at BD5). **e**, for Omicron wave subgroup, the distribution of anti-D614G nAb titers (light red dots) and anti-BA.1 titers at BD8 (dark red dots), among individuals who had one prior SARS-CoV-2 infection before BD8. Each dot represents one individual, with two measurements of the same individual connected through a gray line. **f**, for the Omicron wave subgroup, the distribution of the anti-D614G nAb titer at BD5, stratified by individuals who were infected during the Omicron wave (solid bar) vs those who were not infected (dashed bar). Independent samples t-test (two-sided) is used to determine the statistical significance (p-value reported on the legend) of difference between the GMT of the two groups. **g**, same as **f** but for anti-BA.1 nAb titers at BD8. **d**, same as **f** but for Δ*nAb*^*E*^ (defined as the difference between anti-BA.1 and anti-D614G titers at BD8).

Similarly, for the Omicron wave, we focused on a subgroup of 535 participants from 184 households who had only one prior SARS-CoV-2 infection (vaccinated and repeatedly infected individuals were removed from the analysis) or remained naïve just before the Omicron wave (see Table 1 and Extended Data Fig. 2 for a description of participants and Fig. 1 for the timing of the Omicron wave). Prior SARS-CoV-2 infection was ascertained in a similar fashion as for the Delta wave (i.e., positivity by anti-nucleocapsid assay and/or rRT-PCR for the first 8 blood draws). Infections during the Omicron wave were inferred based on the anti-nucleocapsid antibody level of two pre- and one post-Omicron wave serum samples, as previously described ^30^. Two thirds, or 67% (359/535), of participants included in the Omicron BA.1/2 wave analysis were infected by these variants, with attack rates of 77% (149/193) and 61% (210/342) for naïve and previously infected individuals, respectively.

To evaluate nAbs as CoP in the context of Omicron’s extensive immune escape, we measured both the anti-D614G nAb titers and anti-Omicron BA.1 nAb titers for serum samples collected at blood draw 8 (the blood draw taken shortly before the onset of the Omicron wave, referred to as BD8 hereafter). Given that none of the participants had been infected by Omicron prior to BD8, the anti-Omicron BA.1 neutralizing activity at this time point originated from cross-reactive antibodies elicited by prior variant infections. Thus, the difference between anti-D614G and anti-BA.1 nAb titers at BD8 represents the quantity of anti-D614G nAbs that failed to recognize mutated epitopes on Omicron BA.1, resulting in a lack of neutralizing function against Omicron BA.1. For the remainder of the manuscript, we will use Δ*nAb*^*E*^ to represent the quantity of antibodies able to neutralize D614G but not Omicron BA.1 due to mutations in the Omicron spike. Similarly to the Delta wave subgroup, a significant proportion of previously infected individuals in the Omicron wave subgroup exhibited anti-D614G and anti-Omicron nAb titers below the detection threshold at BD8 (Extended Data Table 1). The absence of detectable nAbs was also more pronounced when the variant causing prior exposure and the spike used in the neutralization assay were mismatched. (Extended Data Table 1). Roche Elecsys anti-nucleocapsid assay remained robust in detecting prior infection ^33^, despite low nAb titer level (Extended Data Table 1).

Fig. 2e shows the anti-D614G and the anti-BA.1 nAb titers at BD8 for participants included in the Omicron wave analysis. The nAb GMT against D614G was 122 (95% CI 103 – 145) and 30 (95% CI 27 – 34) for antibodies that could neutralize BA.1, representing an average 4.01 (95% CI 3.53 – 4.58) -fold reduction attributed to the immune evasive properties of Omicron. The anti-D614G and anti-BA.1 nAb titers (in log scale) at BD 8 were modestly correlated (Pearson correlation coefficient 0.64, p<0.0001). Comparing the nAb titers between individuals who were infected during the Omicron wave vs. those who were not infected, we did not find significant differences in GMT levels for anti-D614G nAb, anti-BA.1 nAb, or Δ*nAb*^*E*^ (Fig. f-h). However, it is worth noting that the point estimates of GMTs were higher for uninfected individuals compared to infected individuals across all three measurements.

### Pre-exposure nAb titer as CoP against variant infection

We conducted mediation analyses in a household transmission modelling framework to investigate how nAb titers against SARS-CoV-2 variants at the onset of a SARS-CoV-2 wave mediate the risk of infection during the corresponding epidemic wave ^34,35^. Specifically, following the causal inference framework proposed by Halloran and Struchiner ^36^, we introduced SARS-CoV-2 transmission probabilities as causal parameters, representing either the risk of acquiring infection from the general community or the per-contact transmission risk within the household. Transmission probabilities were dependent on an individual’s prior infection history, the level of pre-existing nAb titers (mediators), and other confounding factors (age, gender, comorbidities). We fitted a chain-binomial household transmission model, parametrized by the transmission probabilities, to the infection outcomes of the Delta and Omicron waves among all subgroup participants and evaluated how the level of nAb titers mediated SARS-CoV-2 transmission probability. The details of the mediation analysis are described in the Methods.

For the Delta wave mediation analysis, we considered anti-D614G nAb titer at BD5 as candidate mediator of protection and the quantity of antibodies that had waned from peak (Δ*nAb*^*W*^) as putative negative control (i.e., we hypothesize that antibodies lost due to waning could not conceivably contribute to protection). For the Omicron wave, we considered anti-BA.1 nAb titer at BD8 as candidate mediator of protection and the quantity of nAbs that escape Omicron neutralization (Δ*nAb*^*E*^) at BD8 as putative negative control. We used the term “direct effect” from the causal inference framework to refer to the effect of exposure (prior infection) on the outcome (repeat infection during the Delta or Omicron wave) absent the mediators (nAb titers). Conversely, the term “indirect effect” represents the effect of exposure (prior infection) on the outcome (repeat infection) that operates through the mediators (nAb titers). We estimated both the direct effect of prior infection and effects mediated through specific nAb titers against serologically confirmed SARS-CoV-2 infections. We report the estimates of the mediation analysis for both Delta and Omicron wave in Table 2. For the ease of interpretation, we then translate the estimated odds ratios into risk reductions (1 – odds ratio), along with other estimates in causal diagrams depicted in Fig. 3.

**Table 2:**
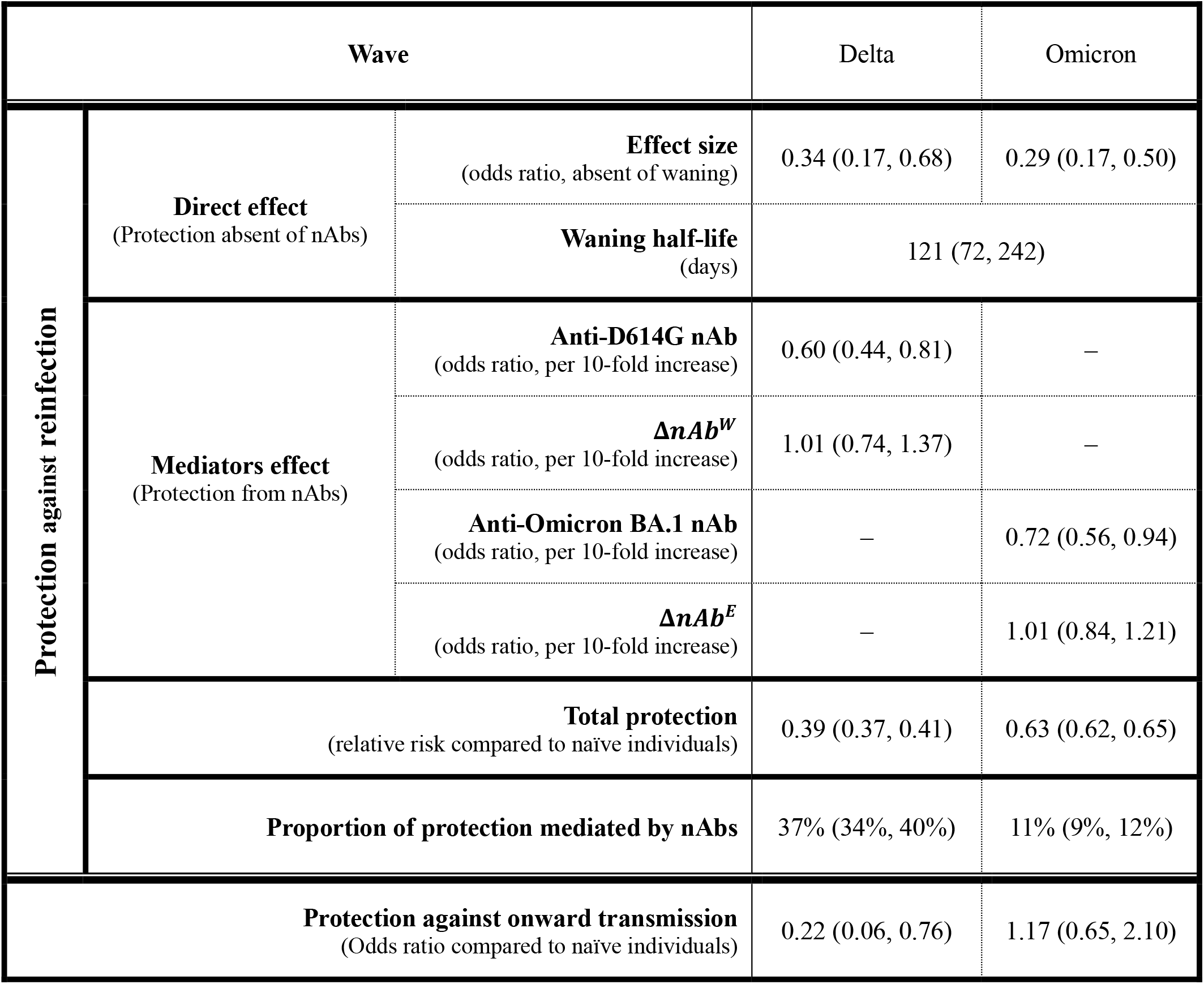
Mediation analysis for nAbs as CoPs against Delta and Omicron wave infections, with a waning model for direct effect. Average and 95% CIs are provided for each of the model parameters. Δ*nAb*^*W*^: the quantity of anti-D614G nAbs waned from peak level to that at BD5. Δ*nAb*^*E*^: the quantity of antibodies that can neutralize D614G but fail to neutralize Omicron BA.1 at BD8 due to Omicron’s immune escape.

**Fig. 3:**
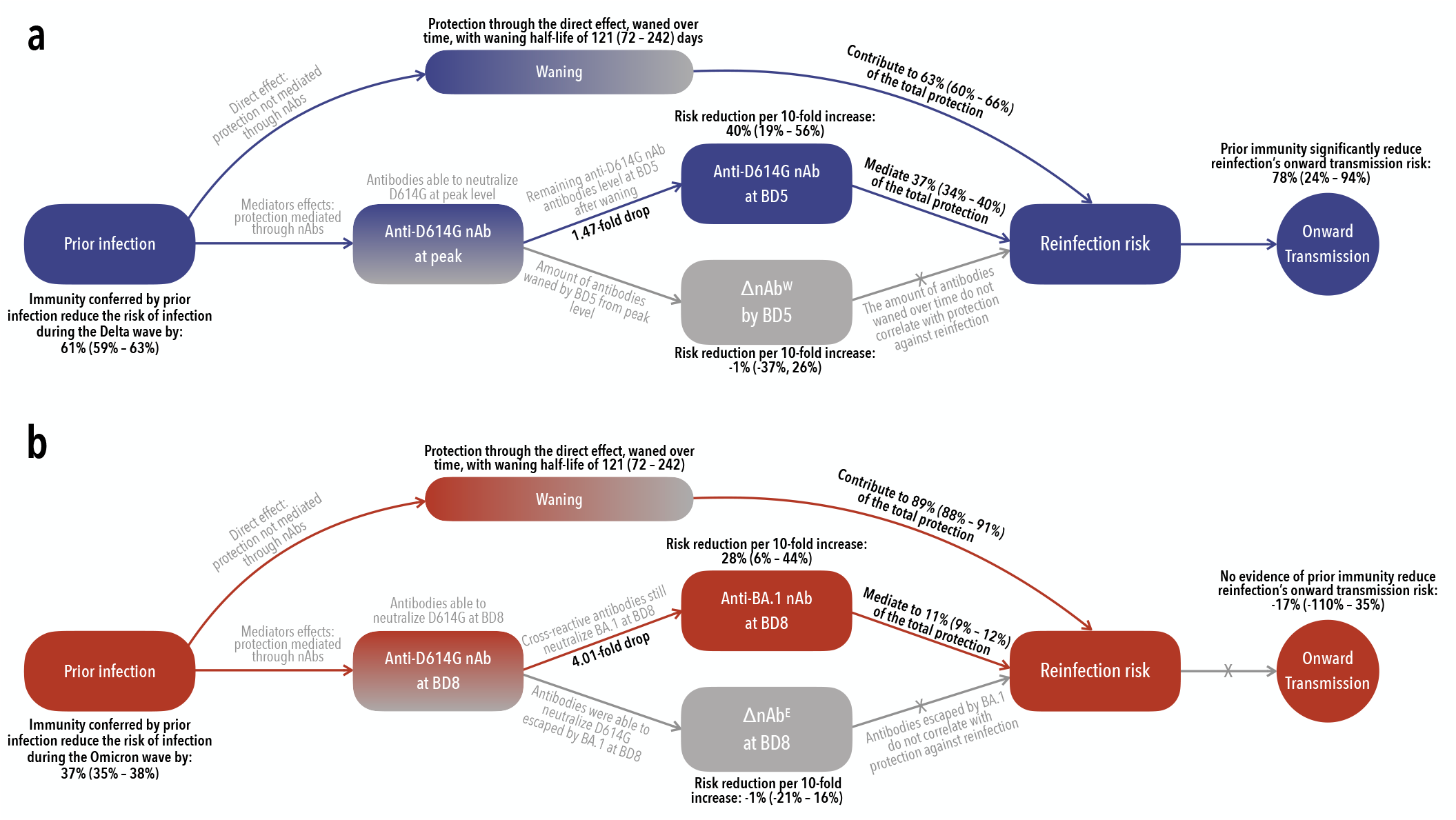
Causal diagrams for the mediation analyses. **a:** Causal diagram of the Delta wave mediation analysis showing the hypothesized relationship between prior immunity (induced by prior SARS-CoV-2 infection) and SARS-CoV-2 infection (outcome of interest) during Delta wave. The mediators of interest are anti-D614G nAbs at BD5 and Δ*nAb*^*W*^ (the quantity of anti-D614G nAbs waned from peak level to that at BD5). The direct effect represents protection operating through immune mechanisms other than the mediators of interest. We hypothesize that the direct effect could wane over time since the initial immune exposure. For the prospective cohort data, both mediator-outcome confounding and exposure-outcome confounding factors need to be adjusted for in the mediation analysis, as the immune exposure (prior SARS-CoV-2 infection) was not randomly assigned (unlike SARS-CoV-2 randomized-control vaccine trials where vaccination was randomly assigned to the participants). Furthermore, cohort participants may experience heterogenous levels of SARS-CoV-2 exposure due to different intensity SARS-CoV-2 transmission in their household settings. We adjust this by embedding the mediation analysis in a mechanistic household transmission model (detailed in Methods). We also look at the impact of prior immunity on the reduction of onward transmission, conditional on the failure of preventing reinfection. The estimates of the Delta wave mediation analysis are presented in Table 2. **b:** Same as a but for the Omicron wave analysis. The mediators of interest are anti-BA.1 nAbs at BD8 and Δ*nAb*^*E*^ (the quantity of antibodies that can neutralize D614G but fail to neutralize Omicron BA.1 at BD8 due to Omicron’s immune escape.). The estimates of the Omicron wave mediation analysis are presented in Table 2.

Our findings indicate that immunity derived from prior infection, overall, reduced the risk of contracting a Delta wave infection by 61% (95%CI: 59% – 63%) (Fig 3a). Notably, nAbs represented an important mediator of this overall protection: for every 10-fold increase in the anti-D614G nAb titers at BD5, the risk of infection decreased by 40% (95% CI 19% – 56%). In contrast, the decline in nAbs from peak levels to BD5 (Δ*nAb*^*W*^) showed no contribution to the overall protection, with a risk reduction per 10-fold increase of -1% (95%CI: -21% – 16%). This result indicated that waning of neutralizing antibodies results in waning of protection, in agreement with our hypothesis. Furthermore, we estimated that the protection mediated through anti-D614G nAbs at BD5 accounted for 37% (95% CI: 34% - 40%) of the overall protection derived from prior infection, suggesting that over half of the protection against Delta was not mediated by serum nAbs against D614G. Lastly, our analysis indicated that individuals reinfected with the Delta variant were 78% (95% CI: 24% – 94%) less likely to transmit the infection to other household members compared to those who experienced primary infections (Fig. 3a). This finding suggested that even in cases where prior immunity is not sufficient to block reinfection with the Delta variant, infection-induced immunity still offered sizable mitigation against onward transmission.

The causal diagram depicting the mediation analysis for the Omicron wave is illustrated in Fig. 3b. Our findings indicate that, overall, prior infection-derived immunity resulted in a 37% (95%CI: 35% – 38%) reduction in the risk of contracting an Omicron wave infection, a notably lower effect compared to that of the Delta wave. We observed that, anti-Omicron nAbs at BD8 significantly mediated protection against the Omicron BA.1/2 variants: for every 10-fold increase in anti-Omicron nAb titers, the risk of Omicron BA.1/2 infection decreased by 28% (95% CI: 6% – 44%). Conversely, antibodies unable to neutralize Omicron due to immune escape (Δ*nAb*^*E*^) did not mediate protection against Omicron BA.1/2 infection, with risk reduction of -1% (95% CI: -21% – 16%) per 10-fold titer increase. Furthermore, we estimated that the protection mediated through anti-BA.1 nAbs at BD8 accounted for only 11% (95% CI: 9% - 12%) of the total protection conferred by prior exposure. This, coupled with the observation that Omicron BA.1 caused an average of 4.01-fold drop in nAb titers (Fig. 2e), underscores Omicron BA.1/2’s ability to evade host protective immunity mediated through nAbs. Additionally, in contrast to the Delta wave, individuals reinfected with the Omicron variant were as likely to transmit the infection to other household members compared to those who experienced primary infections (risk reduction: -17%, (95% CI: -110% – 35%)). These observations suggested that Omicron not only evaded prior immunity’s protection against acquisition of infection but also escaped protection against onward transmission.

Although neutralizing titers measured at BD5 and BD8 offered a temporally proximate evaluation of protective immunity preceding the onset of the Delta and Omicron waves, we could not identify the immune mediators responsible for the direct effects of prior immunity (i.e., the fraction of protection that was not mediated by nAbs) due to lack of additional serum biomarkers. We could however estimate the potential for these direct effects to wane over time. To do so, we modeled an exponential decline for the direct effect based on the time elapsed since prior infection and jointly estimated the duration of protection for both the Delta and Omicron waves’ analysis. We found that protection not mediated by nAbs decreased with time, with a waning half-life of 121 (95%CI: 72 – 242) days (Fig. 3, Table 2). After adjusting for waning, the effect sizes of protection from direct effects were similar for both variants, with odds ratios of acquiring infection (compared to naïve individuals) of 0.34 (95% CI: 0.17, 0.68) and 0.29 (95% CI: 0.17, 0.50) for the Delta and Omicron wave, respectively, in the absence of waning (Table 2). These results suggested that, while Omicron escaped pre-existing neutralizing antibodies, protection from other immune effectors was preserved against this variant. The waning half-life of protection not mediated by nAbs was estimated at approximately 4 months in our study, comparable to the reported waning timescale of T-cell immunity ^37,38^. Several sensitivity analyses demonstrating the robustness of the findings of the mediation analysis were reported in the Methods.

## Discussion

In this cohort of unvaccinated individuals, we found that nAb titers immediately before the onset of the Delta wave (i.e., anti-D614G nAb level at BD5) correlated with protection against Delta wave infections. Moreover, we demonstrated that nAb titers lost over time due to waning (i.e., *ΔnAb*^*W*^) were not associated with protection, aligning with the expectation that waning of nAbs in serum corresponds to waning of clinical protection. For the Omicron wave subgroup, we further investigated the impact of immune escape against protection mediated through nAbs. We found that only anti-Omicron BA.1 nAbs correlated with protection against infection during the Omicron BA.1/2 wave, whereas anti-D614G nAbs that were unable to neutralize Omicron BA.1 due to spike escape mutations did not protect. This indicated that antibodies capable of neutralizing D614G but not Omicron BA.1 *in vitro* translates to a diminished protection against Omicron BA.1/2 infection among PHIRST-C participants. The identification of variant-neutralizing antibodies derived from infection-induced immunity as CoPs against infections for both Delta and Omicron variants aligns with findings from studies on variant-specific correlates for vaccine-induced or hybrid immunity ^21–24^. Considering that antibody-mediated protection against acquisition of infection likely operates at the mucosal site rather than in serum, it is interesting that serum antibodies levels can anticipate protection ^26^. In a recent analysis of the data from the COVE trial, Zhang et. al. further demonstrated that boosting of nAb titers against Omicron by a third dose of mRNA-1273 vaccine, afforded additional protection against Omicron compared to individuals who only received 2 doses of the mRNA-1273 vaccine. ^22^ Collectively, these empirical data lend support for using nAbs against circulating variant as immuno-bridging markers for periodic vaccine updates.

While a comprehensive understanding of the role of nAbs in SARS-CoV-2 protection is important, a key finding of our study is that nAb titers did not fully mediate protection conferred by prior infection. In the case of the Delta wave subgroup, we estimate that anti-D614G nAbs mediate 37% of protection, a proportion comparable to vaccine-induced nAbs ^15^. In contrast, for the Omicron BA.1/2 wave subgroup, anti-Omicron BA.1 nAbs are estimated to mediate only 11% of protection, which was substantially lower than that observed for the Delta wave. This low percentage of protection mediated by nAbs for the Omicron BA.1/2 wave could be attributed to the highly immune evasive nature of Omicron against neutralizing activity. Omicron effectively rendered a significant proportion of serum anti-D614G nAbs nonfunctional against Omicron. The large proportion of overall protection that was not mediated by nAbs could be explained by a variety of immune mechanisms, including the Fc-effector function of binding antibodies, and T-cell functions, both of which are resilient against mutations in VOCs ^14,39^. Additionally, SARS-CoV-2 initially infects and predominantly transmits through the upper respiratory tract. Mucosal immunity in the upper respiratory tract therefore likely plays a key role in preventing SARS-CoV-2 infection, and may not be fully represented by immune markers in serum ^40^. Our study validates the use of serum nAbs as CoP against reinfection but also suggests potential important roles for other candidate functions that could act as “co-correlates” of protection ^41^. This is particularly important because these mechanisms may be more broadly cross-protective against future variants than neutralizing antibodies. Future CoP analyses incorporating measurements of T-cell immunity and non-neutralizing antibody functions, ideally at the mucosal site, could potentially disentangle these important protective mechanisms and inform design of next generation vaccines ^26,42–44^.

Our study has several limitations. Firstly, the vaccination rate in the PHIRST-C cohort was low at the time of the analysis; with <20% participants fully vaccinated prior to the Omicron BA.1/2 wave (thus excluded from our analysis). Consequently, we lacked sufficient statistical power to assess CoPs for vaccine-induced (or hybrid) immunity and compare with our findings for infection-induced immunity in the same cohort. Secondly, we focused on SARS-CoV-2 infections that were ascertained by seroconversion or amnestic boosting of the anti-nucleocapsid antibodies. However, not all PCR-positive SARS-CoV-2 infections led to systemic antibody response ^30,45,46^. Thus, our CoP analysis does not account for protection against abortive or transient infections that do not lead to systemic antibody responses. We also could not evaluate CoPs against symptomatic cases, as there was no systemic monitoring of SARS-CoV-2 symptoms for the cohort population during the Omicron BA.1/2 wave. Further, severe outcomes (hospitalizations and deaths) due to SARS-CoV-2 infections were rare throughout the course of the PHIRST-C study and evaluation of protection against those outcomes is under-powered. Understanding protection against severe outcomes is important from both clinical and public health prospective, thus warranting further studies. Thirdly, the strains of antigens used in neutralization assay were not perfectly matched to the circulating variants in the CoP analysis. For the Delta wave analysis, we evaluated anti-D614G antibody titers (rather than anti-Delta titers). Although Delta is not as immune evasive as Omicron with respect to D614G, there are substitutions on the spike of Delta (i.e., L452R, T478K) that are linked to moderate antigenic escape ^47,48^. In addition, though infections were predominantly caused by the Delta variant during the Delta wave epidemic, other variants also circulated at low levels during the same time period, including Alpha and C.1.2 ^30^. Similarly, genomic surveillance revealed that while Omicron BA.1 accounted for the majority of infections during the Omicron wave, Omicron BA.2 also co-circulated, with potential antigenic spike substitutions (e.g., T376A, D405N, R408S) that were not present in BA.1 ^30,48,49^. Thus, using a BA.1-specific neutralizing assay may introduce bias in our CoP analysis, particularly against Omicron BA.2. Lastly, we only measured serum antibodies, but did not have any information on antibody response at the mucosal site or on cell-mediated immunity. While serum IgG nAbs may transudate into the nasal mucosa and thereby play a role in protection, the contribution of locally produced nasal IgA nAb remains to be investigated.

Moving forward, future works focusing on understanding how protective immunity accumulates through repeated infections, vaccinations, and hybrid immunity, and identifying a suite of predictive markers of protection reflecting different arms of immune responses, are key to anticipating long-term SARS-CoV-2 burden, optimizing vaccine boosters, and designing next generation SARS-CoV-2 vaccines.

## Acknowledgment

We are thankful to all the participants who kindly agreed to take part in the study, as well as to the PHIRST group. We are grateful to Professor Benjamin J. Cowling and Professor M. Elizabeth Halloran for their insightful feedback on the paper. We deeply appreciate the three anonymous reviewers for their constructive review of the paper. The findings and conclusions in this report are those of the authors and do not necessarily represent the official position of the NIH or the U.S. Centers for Disease Control and Prevention. **Funding:** This work was supported by the National Institute for Communicable Diseases of the National Health Laboratory Service and the U.S. Centers for Disease Control and Prevention [cooperative agreement number: 6 U01IP001048] and Wellcome Trust (grant number 221003/Z/20/Z) in collaboration with the Foreign, Commonwealth and Development Office, United Kingdom. PLM and JNB are supported by the Bill and Melinda Gates Foundation through the Global Immunology and Immune Sequencing for Epidemic Response (GIISER) program (INV-030570) and receive funding from the Wellcome Trust (226137/Z/22/Z). PLM is supported by the South African Research Chairs Initiative of the Department of Science and Innovation and National Research Foundation of South Africa and the SA Medical Research Council SHIP program.

## Author Contributions Statement

KS, JNB, ST, JK, AvG, MLM, NW, JM, NAM, KK, LL, CV, PLM, CC designed the experiments. JNB, CC, JK, PLM and ST accessed and verified the underlying data. JNB, ST, JK, VSM, QM, HK, AvG, MLM, NW, JM, MC, NAM, KK, LL, JdT, TM, PLM, CC collected the data and performed laboratory experiments. KS, JNB, ST, JK, AvG, MLM, NW, JM, MC, NAM, KK, LL, JdT, TM, CV, and CC analyzed the data and interpreted the results. KS, JNB, CV, PLM, and CC drafted the manuscript. All authors critically reviewed the article. All authors had access to all the data reported in the study.

## Consortia Authorship

The PHIRST-C group: Jinal N. Bhiman^2,3^, Amelia Buys^4^, Maimuna Carrim^4,7^, Cheryl Cohen^4,5^, Linda de Gouveia^4^, Mignon du Plessis^4,7^, Jacques du Toit^11^, Francesc Xavier Gómez-Olivé^11^, Kathleen Kahn^11^, Kgaugelo Patricia Kgasago^9^, Jackie Kleynhans^4,5^, Retshidisitswe Kotane^4^, Limakatso Lebina^9^, Neil A Martinson^9,10^, Meredith L McMorrow^6,8^, Tumelo Moloantoa^4^, Jocelyn Moyes^4,5^, Stefano Tempia^4,5,6^, Stephen Tollman^11^, Anne von Gottberg^4,7^, Floidy Wafawanaka^11^, Nicole Wolter^4,7^

## Methods

### Ethics Statement

The PHIRST-C protocol was approved by the University of Witwatersrand Human Research Ethics Committee (Reference 150808) and the U.S. Centers for Disease Control and Prevention’s Institutional Review Board relied on the local review (#6840). The protocol was registered on clinicaltrials.gov on 6 August 2015 and updated on 30 December 2020 (https://clinicaltrials.gov/ct2/show/NCT02519803). Participants receive grocery store vouchers of ZAR50 (USD 3) per visit to compensate for time required for specimen collection and interview. All participants provided written informed consent for study participation. For minors, consent was obtained from the parent or guardian.

### Inferring Delta and Omicron wave infections based on longitudinal serum samples

We have previously described the serologic inference method for SARS-CoV-2 infections among the PHIRST-C cohort participants during the Delta wave (3^rd^ SARS-CoV-2 wave) and the Omicron wave (4^th^ SARS-CoV-2 wave) ^30^. To briefly summarize, ascertainment of Delta wave infections was based on the serial serologic readout of blood draws 5 and 6 (both before the Delta wave, figure 1A-B), and 8 (post Delta wave), measured by the Roche Elecsys Anti-SARS-CoV-2 nucleocapsid assay ^33^. The participants’ serologic trajectories were then grouped into 13 categories of distinct serum antibody patterns, reflecting the rise, waning, and/or amnestic boosting of anti-nucleocapsid antibody levels. Because the Delta wave was also covered by intense virologic sampling with twice-weekly nasopharyngeal swab collection, we grouped the 13 serologic categories into indicators of either presence or absence of SARS-CoV-2 infection to achieve the highest concordance with rRT-PCR-confirmed Delta infections. The Omicron wave was not covered by the intense PCR testing; however, the timing of blood draws 8, 9, and 10 with respect to the Omicron wave is similar to that of blood draws 5, 6, and 8 with respect to the Delta wave (figure 1A-B). We thus apply the same classification method of serial serologic trajectories defined by blood draws 8, 9, and 10 to infer SARS-CoV-2 infections during the Omicron BA.1/2 wave.

### Laboratory Methods

#### Serum nAb titers against SARS-CoV-2 D614G and BA.1 variants (Lentiviral Pseudovirus Production and Neutralization Assay)

Virus production and pseudovirus neutralization assays were done as previously described ^50^. Briefly, 293T/ACE2.MF cells modified to overexpress human ACE2 (kindly provided by M. Farzan (Scripps Research)) were cultured in DMEM (Gibco BRL Life Technologies) containing 10% heat-inactivated serum (FBS) and 3 μg ml^−1^ puromycin at 37 °C, 5% CO_2_. Cell monolayers were disrupted at confluency by treatment with 0.25% trypsin in 1 mM EDTA (Gibco BRL Life Technologies). The SARS-CoV-2, Wuhan-1 spike, cloned into pCDNA3.1 was mutated using the QuikChange Lightning Site-Directed Mutagenesis kit (Agilent Technologies) and NEBuilder HiFi DNA Assembly Master Mix (NEB) to include D614G (wild-type) or lineage defining mutations for Delta (T19R, 156-157del, R158G, L452R, T478K, D614G, P681R and D950N) and), Omicron BA.1 (A67V, 69-70del, T95I, G142D, 143-145del, 211del, L212I, 214EPE, G339D, S371L, S373P, S375F, K417N, N440K, G446S, S477N, T478K, E484A, Q493R, G496S, Q498R, N501Y, Y505H, T547K, D614G, H655Y, N679K, P681H, N764K, D796Y, N856K, Q954H, N969K, L981F), Omicron BA.2 (T19I, L24S, 25-27del, G142D, V213G, G339D, S371F, S373P, S375F, T376A, D405N, R408S, K417N, N440K, S477N, T478K, E484A, Q493R, Q498R, N501Y, Y505H, D614G, H655Y, N679K, P681H, N764K, D796Y, Q954H, N969K). Pseudoviruses were produced by co-transfection in 293T/17 cells with a lentiviral backbone (HIV-1 pNL4.luc encoding the firefly luciferase gene) and either of the full-length SARS-CoV-2 spike plasmids with PEIMAX (Polysciences). Culture supernatants were clarified of cells by a 0.45μM filter and stored at −70 °C. Plasma samples were heat-inactivated and clarified by centrifugation. Pseudovirus and serially diluted plasma/sera were incubated for 1 h at 37 °C, 5% CO_2_. Cells were added at 1 × 10^4^ cells per well after 72 h of incubation at 37 °C, 5% CO_2_, luminescence was measured using PerkinElmer Life Sciences Model Victor X luminometer. Neutralization was measured as described by a reduction in luciferase gene expression after single-round infection of 293T/ACE2.MF cells with spike-pseudotyped viruses. Titers were calculated as the reciprocal plasma dilution (ID_50_) causing 50% reduction of relative light units.

Noting that we measured neutralization titer using a lentiviral-backboned pseudovirus neutralization assay. A systematic review of Omicron neutralization data showed that pseudovirus neutralization assays tend to report higher neutralizing titers compared to live-virus assays. The titer drops from wild type to Omicron also tend to be less pronounced for pseudovirus platforms, suggesting the pseudovirus assay may underestimate Omicron’s capability to escape neutralization

#### sars-CoV-2 spike enzyme-linked immunosorbent assay (ELISA)

For ELISA, Hexapro SARS-CoV-2 full spike protein with the D614G substitution was expressed in Human Embryonic Kidney (HEK) 293F suspension cells by transfecting the cells with the respective expression plasmid. After incubating for 6 days at 37°C, proteins were first purified using a nickel resin followed by size exclusion chromatography. Relevant fractions were collected and frozen at -80°C until use. Two µg/mL of D614G spike protein was used to coat 96-well, high-binding plates (Corning) and incubated overnight at 4°C. The plates were incubated in a blocking buffer consisting of 1x PBS, 5% skimmed milk powder, 0.05% Tween 20. Plasma samples were diluted to 1:100 starting dilution in a blocking buffer and added to the plates. IgG secondary antibody (Merck) was diluted to 1:3000 in blocking buffer and added to the plates followed by TMB substrate (Thermofisher Scientific). Upon stopping the reaction with 1 M H2SO4, optical density (OD) was measured at 450 nm. The monoclonal antibodies (mAbs) CR3022 and Palivizumab were used as the positive and negative controls respectively.

### Statistical Analysis

#### Mediation analyses and household transmission model fitted to observed infections in the cohort

Here we blend concepts from causal inference and infectious disease transmission models. The no interference assumption in causal inference stipulates that the outcome of an individual does not depend on the outcome of others, which is often violated in infectious disease dynamics ^36,52,53^. This is because the spread of infectious diseases requires pathogens to be transmitted from one host to another. In other words, the infection outcome of one individual inherently depends on the infection outcome of others, and this is particularly pronounced in a household setting ^36^. The “dependent happening” nature of infectious disease dynamics violates the no interference assumption. As a result, the traditional regression approach for causal inference analysis cannot be applied to infectious disease outcomes among individuals who can in theory transmit the disease from one to another. To overcome this, Halloran and Struchiner. ^36^ introduced the probability of infection conditional on exposure to already infected individuals (transmission probability), as the causal parameter. Using this proposed framework, we can investigate how the presence/absence of pre-existing immunity along with the immunologic marker of interest could modulate probability of infection, after adjusting for levels of exposure to the infectious source(s). The corresponding causal inference framework requires modelling the transmission process explicitly. Under this framework, we conduct mediation analyses to investigate how nAb titers against variants at the start of a SARS-CoV-2 wave correlate with SARS-CoV-2 transmission risk, using the Delta and Omicron BA.1/2 waves as examples ^34,35^. We focus on the Delta and Omicron subgroup participants who have had a single or no prior infection, and fit a chain-binoal model to their infection outcomes during the corresponding Delta/Omicron wave ^54^. Specifically, we introduce the causal parameters:

- 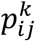: the per-contact SARS-CoV-2 household transmission probability from infected individual *i* to individual *j* in household *k*.
- 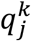: the overall probability of acquiring SARS-CoV-2 infection from outside the household by individual *j* of household *k* (probability of infection from the community).

We use *e*_*j*_ to indicate individual *j*’s prior SARS-CoV-2 infection history, with *e*_*j*_ = 0 representing no prior infection reported before the start of Delta/Omicron BA.1/2 and *e*_*j*_ = 1 representing one prior infection by the start of Delta/Omicron BA.1/2 wave. A prior SARS-CoV-2 infection (*e*_*j*_ = 1) would induce immunologic responses, measured by a set of immune markers (i.e., candidate mediators) {*m*_*j*_|*e*_*j*_ = 1} (e.g., nAb titers level). Then the household transmission probability 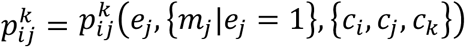 can be expressed as a function of prior infection status *e*_*j*_, immunologic mediators of SARS-CoV-2 transmission probability {*m*_*j*_ |*e*_*j*_ = 1} and additional adjustment terms {*c*_*i*_, *c*_*j*_, *c*_*k*_}, representing a set of potential confounding factors of individual *i*, individual *j*, and household *k* (eg, age of the donor and/or recipient, comorbidities, household size, etc). Similarly, the community infection probability 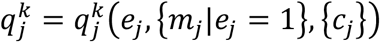 can be expressed as a function of individual *j* ‘s prior exposure history *e*_*j*_, immunological markers {*m*_*j*_ |*e*_*j*_ = 1}, and additional adjustment terms {*c*_*j*_}, representing a set of potential confounding factors of individual *j* (e.g., age or comorbidities).

The causal diagram of the mediation analysis framework is shown in Fig. 3. We fit a household transmission model to the imputed household transmission chains based on an Expectation-maximization (EM) algorithm (detailed in Section 4). Specifically, for the Delta/Omicron BA.1/2 wave, if we look into a specific household *k* of size *N*, there are a total of *n* individuals infected belonging to *L* distinct chains of transmission due to *L* independent introductions of SARS-CoV-2 into the household. The uninfected individuals are *N* − *n*. We denote 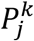 the likelihood of any individual *j* of household *k* having the observed infection status over the Delta/Omicron BA.1/2 wave (i.e., either infected or not) in a particular realization of the model. There are a few scenarios to write down 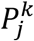:

- Within a given transmission chain *l* ∈ *L*, the initial generation 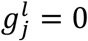 always has an individual *j* acquiring infection from the general community (outside the household *k*). Thus, the probability of individual *j* being infected is 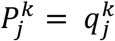 if *j* is the first individual to be infected in the chain.
- For infected individual *j* in the first generation of transmission chain *l*, i.e.,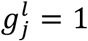, this individual would have to escape infection risk from the general community but get infected by the infected household member of 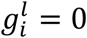. Thus, the probability of individual *j* being infected can be written as 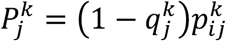.
- For infected individual *j* in transmission chain *l* with generation greater than 1, i.e., *g*_*I*_ > 1, this individual has escaped infection risk from the general community as well as infected individuals *i* two generations away 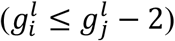 but got infected by an infector *i*^′^ of *j*’s previous generation on the same transmission chain *l*. Thus, the probability of individual *j* being infected can be written as 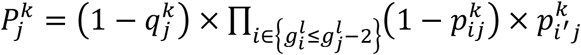.
- For uninfected individual *j* within household *k*, this individual has escaped infection risk from the general community as well as all the *n* infected individuals within the same household. Thus, the probability of individual *j* remaining uninfected can be written as 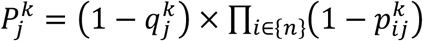.

Then, within household *k* of size *N*, we can express the likelihood of transmission chain *l* as 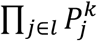; the likelihood of observing all infections within *k* can be expressed as 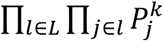; the likelihood of observing *N* − *n* uninfected individuals can be expressed as 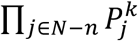. Putting these together, the likelihood of observing one realization of the imputed (details of the EM imputation method described in the next section) households’ transmission trees for Delta/Omicron wave can be expressed as:

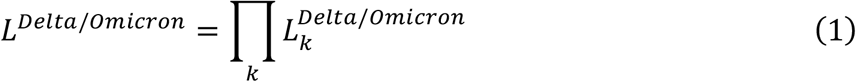

Where the likelihood of a given household transmission chains configuration 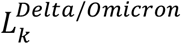 can be expressed as:

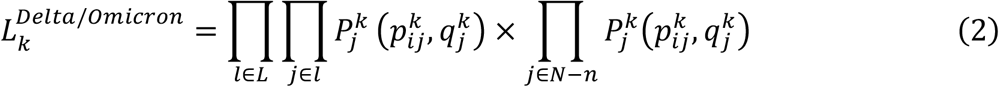

In the remainder of the section, we will consider a few versions of the transmission model with slightly different implementations for 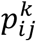 and 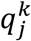.

#### Model 1: waning model for prior exposure with serologically ascertained Delta and Omicron wave infections

This is the transmission model presented in the main analysis of the manuscript (results of the model shown in Table 2. In this model, we consider that protection from prior infection unexplained by nAb titers wanes over time but is not dependent on the variant responsible for prior infection (i.e. prior D614G or Beta infections for the Delta wave analysis, and prior D614G, Beta, or Delta infections for the Omicron wave analysis). Additionally, in this model, both the Delta and Omicron wave infections were ascertained by serology based on approach describe in a prior session in Methods.

More specifically, for the Delta wave, 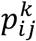 and 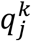 can be expressed as:

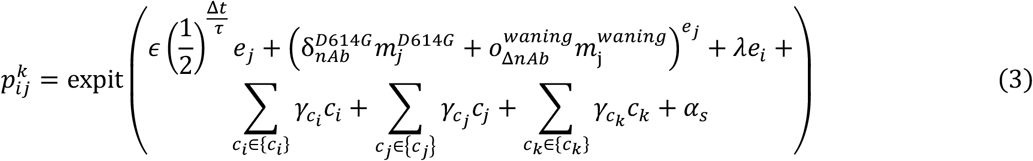

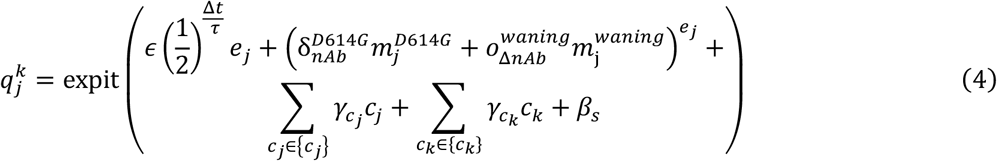

As described before, *e*_*j*_ indicates individual *j*’s prior SARS-CoV-2 infection history, with *e*_*j*_ = 0 representing uninfected individuals at the start of the Delta wave, *e*_*j*_ = 1 representing one prior infection, and *ϵ* representing the effect size of the immune protection by prior infection not mediated through anti-D614G nAbs (direct effect, Table 2). Δ*t* is the elapsed time between prior infection and blood draw 5 (the blood draw taken prior to the Delta wave which we use in this model) and *τ* is the waning half-life of *ϵ* (direct effect, Table 2). 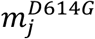 represents the anti-D614G nAb titer at blood draw 5 and 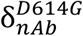 represents the effect size of 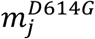 in mediating infection probability 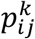 against the Delta wave infection (mediator effect, Table 2) at blood draw 5. While 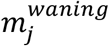 represents the quantity of anti-D614G nAbs waned from peak level (measured as the highest anti-D614G nAb titer level among the first 5 blood draws) to that at BD5 and 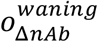 represents the effect size of 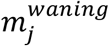 in mediating transmission probability 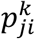 against the Delta wave infection (mediator effect, Table 2) at blood draw 5. Note that the term 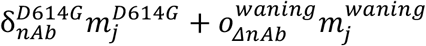 only exists when *e*_*j*_ = 1.

We further evaluate whether breakthrough infections have reduced infectiousness compared to primary infections and may in turn affect 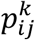. We use *e*_*i*_ to indicate individual *i*’s (the donor) prior SARS-CoV-2 infection history (*e*_*i*_ = 0 means no infection, and *e*_*i*_ = 1 represents one prior infection at the start of Delta wave). Further, *λ* represents the effect size of prior infection (in *i*) in reducing the infectiousness of reinfections.

We also consider confounding factors for donor *i* and recipient *j*, where *c*_*i*_ and 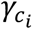 represent infector *i*’s confounding factor (*i*’s age, sex) and effect size, respectively; *c*_*j*_ and 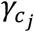 represent *j*’s confounding factor (*j*’s age/sex-specific susceptibility (biology), age/sex- and site-specific susceptibility (behavioral), HIV infection status) and effect size, respectively; *c*_*k*_ and 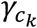 represent household *k*’s confounding factor (household size) and effect size, respectively. Lastly, *α*_*s*_ and *β*_*s*_ are logits of the baseline risks for household and community exposures. All parameters’ effect sizes are measured in the log of odds ratios.

Similarly, for the Omicron BA.1/2 wave, 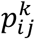 and 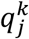 can be expressed a*s*:

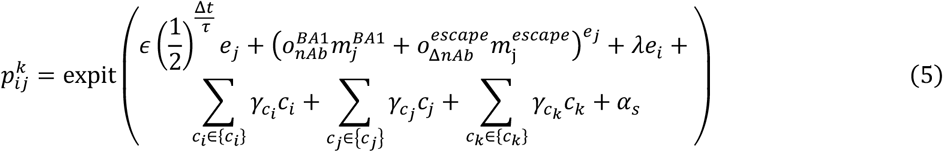

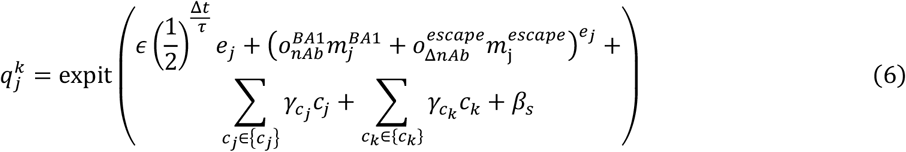

As described before, *e*_*j*_ indicates individual *j*’s prior SARS-CoV-2 infection history, with *e*_*j*_ = 0 representing individual *j* remained naïve to SARS-CoV-2 at the start of Omicron BA.1/2 wave while *e*_*j*_ = 1 representing individual *j* had one prior infection at the start of Omicron BA.1/2 wave and *ϵ* represents the effect size of the immune protection by prior infection not mediated through anti-D614G nAbs (direct effect, Table 2). Δ*t* is the elapsed time between prior infection and blood draw 8 (the blood draw taken prior to the Omicron BA.1/2 wave) and *τ* is the waning half-life of *ϵ* (direct effect, Table 2). Here we consider that parameter *τ* is shared between the Delta and Omicron wave and will be jointly estimated (described in the next session in Methods). 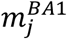 represents the anti-BA1 nAb titer at blood draw 8 and 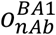 represents the effect size of 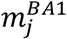 in mediating transmission probability 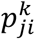 against the Omicron BA.1/2 wave infection (mediator effect, Table 2) at blood draw 8. While 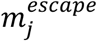 represents the difference in titer from anti-D614G nAb to anti-BA1 nAb at blood draw 8 and 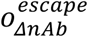 represents the effect size of 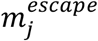 in mediating transmission probability 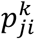 against the Omicron BA.1/2 wave infection (mediator effect, Table 2) at blood draw 8. Note that the term 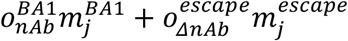 only exists when *e*_*j*_ = 1. All other parameters have the same definition of the Delta wave.

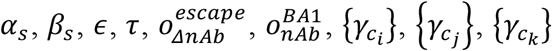 are estimated through maximizing the likelihood function *L* for each of the 100 bootstrapped realizations and bootstrap mean and confidence intervals are calculated for each of the parameters.

### Sensitivity analysis

#### Model 2: Sensitivity analysis considering variant-specific prior exposure for the direct effects

A potential confounding factor in understanding the waning of protection through direct effects is the diversity of prior SARS-CoV-2 exposures, with the dominance of D614G variant in the first wave, Beta variant in the second wave, and Delta variant in the third wave (Fig. 1). The effectiveness of protection may vary depending on the specific variant of prior exposure that induced the immune response at play. We conducted a sensitivity analysis (Model 2) employing a variant-specific model for the direct effects, which accounted for distinct types of SARS-CoV-2 variants conferring prior immunity, instead of considering generic a waning model. Specifically, in Model 2, we considered a more complex version of Model 1, where protection from prior infection depends on the type of infecting variant (i.e. prior D614G or Beta infections for the Delta wave analysis, and prior D614G, Beta, or Delta infections for the Omicron wave analysis). We consider waning in neutralizing titers as in Model 1, but we eliminate waning in the effect of prior infection that is not captured by neutralizing titers. More specifically, for the Delta wave, 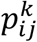 and 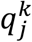 can be expressed as:

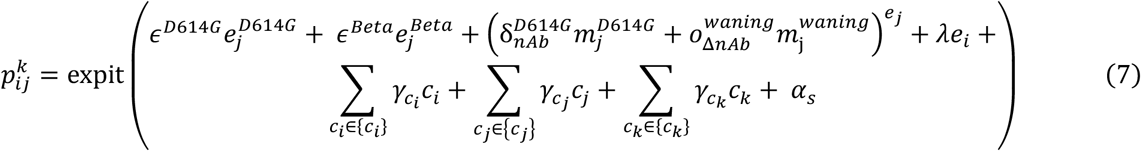

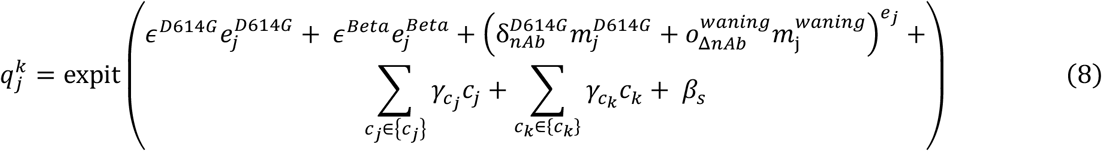

Here, 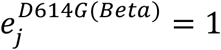 indicates individual *j*, prior to the Delta wave, was infected with D614G (Beta) variant. If 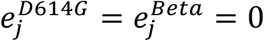, individual *j* was naïve at the beginning of the Delta wave. 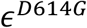 and 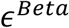 represent the effect size of immune protection by prior D614G and Beta infection not mediated through anti-D614G nAbs, respectively.

For the Omicron wave, 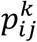 and 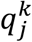 can be expressed as:

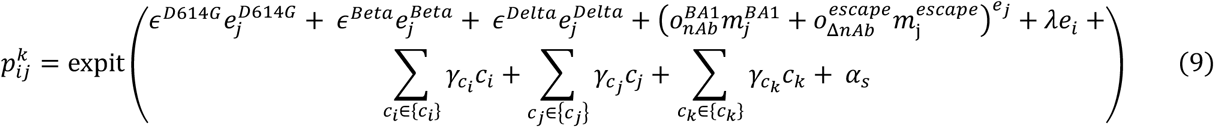

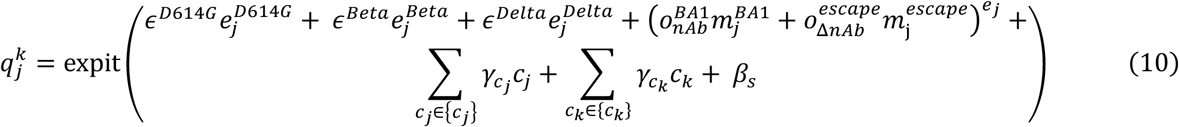

Here, 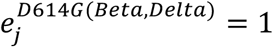 indicates individual *j*, prior to the Omicron wave, was infected with D614G (Beta, Delta) variant. If 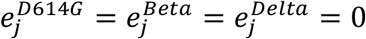, individual *j* was naïve at the beginning of the Omicron wave. 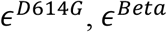 and 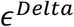 represent the effect size of the immune protection by prior D614G, Beta and Delta infection not mediated through anti-D614G nAbs, respectively.

Additionally, similarly to Model 1, both the Delta and Omicron wave infections were ascertained by serology for Model 2. All other settings of Model 2 were kept the same as Model 1. The results of the Model 2 are presented in Extended Data Table 2.

Our analysis revealed that for both the Delta and Omicron waves, more recent variants conferred stronger protection than earlier variants, albeit with overlapping confidence intervals (Extended Data Table 2). This temporal trend aligns with the expectations of the waning model. Both waning and variant-specific immunity may modulate the direct effects of prior immunity; however, our study lacked sufficient statistical power to jointly estimate the relative contributions of these two factors. Full estimates of this sensitivity analyses are presented in Extended Data Table 2.

#### Model 3: Sensitivity analysis with Delta wave infections ascertained by PCR and/or serology

For Model 1, both the Delta and Omicron wave infection outcomes were inferred using the kinetics of anti-nucleocapsid antibodies from longitudinal serologic sampling, as detailed in previously published studies of the PHIRST-C cohort ^30,32^. This approach for inferring infections based on serology was calibrated against virological evidence of infection during the Delta wave, established through twice-weekly rRT-PCR tests regardless of symptom presentation. However, it should be noted that this calibration did not achieve perfect concordance; the serology approach demonstrated 93% sensitivity and 89% specificity when compared to infections identified by rRT-PCR tests ^30^. To address the uncertainties arising from the imperfect concordance between the two approaches for ascertaining infections, we conducted a sensitivity analysis (Model 3) for the Delta wave, where we considered infections based on rRT-PCR positivity and/or anti-nucleocapsid antibody serology. We identified an additional 17 infections during the Delta wave through this more sensitive infection ascertainment approach, bringing the total number of Delta wave infections to 290. All other settings of Model 3 were kept the same as Model 1. The results of the Model 3 are presented in Extended Data Table 3.

Notably, estimates of the direct and indirect effects of the mediation analysis were comparable between this sensitivity analysis and the main analysis (compare Extended Data Table 3 to Table 1). These findings provide support for the utilization of anti-nucleocapsid serology to ascertain Omicron BA.1/2 wave infections in the studied cohorts, in a period where twice-weekly rRT-PCR testing was not available and confirms the robustness of our CoP analyses.

#### Model 4: D614G spike binding antibodies as mediators of protection

We conducted sensitivity analysis (Model 4) to explore the role of D614G spike binding antibodies (referred to as bAb hereafter), as potential correlates of protection for both Delta and Omicron infections. Employing an in-house enzyme-linked immunosorbent assay (ELISA), we quantified the level of D614G spike bAb based by measuring absorbance at 450nm at an optical density (OD) at peak levels and BD5 (DB8) for the Delta (Omicron) wave analysis (Extended Data Fig. 3). The reduction in binding antibody levels from peak (Δ*bAb*^*W*^) was determined as the difference between OD values at peak and BD5 (BD8) for the Delta (Omicron) wave (Extended Data Fig. 3).

Model 4 builds on Model 2 but replaces nAb titers with D614G spiking binding ELISA readouts as mediators of protection, in order to compare the protection afforded by neutralizing vs binding antibodies. More specifically, for the Delta wave, 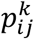 and 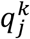 can be expressed as:

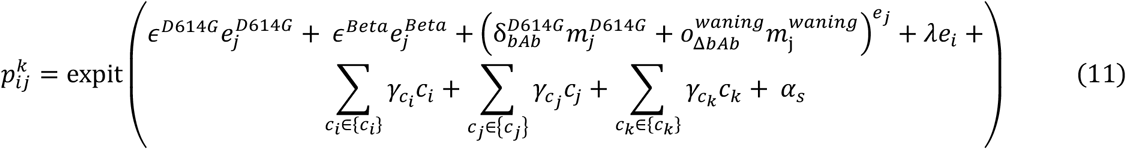

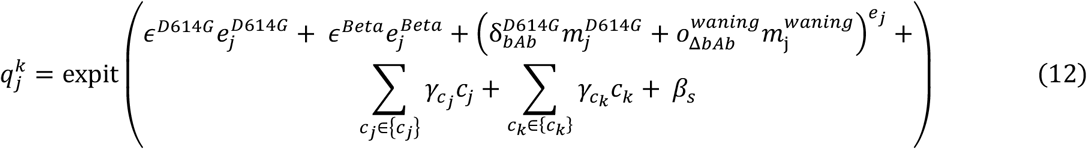

Here, 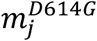 represents the D614G spike binding antibodies ELISA readout at blood draw 5 and 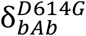 represents the effect size of 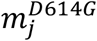 in mediating transmission probability 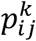 against the Delta wave infection at blood draw 5. Further, 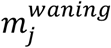 represents the drop from peak D614G spike binding antibodies readout prior to blood draw 5 (measured as the highest D614G spike binding Ab titer level among the first 5 blood draws) to that at blood draw 5 and 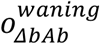 represents the effect size of 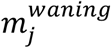 in mediating transmission probability 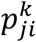 against the Delta wave infection at blood draw 5.

For the Omicron wave, 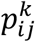 and 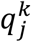 can be expressed as:

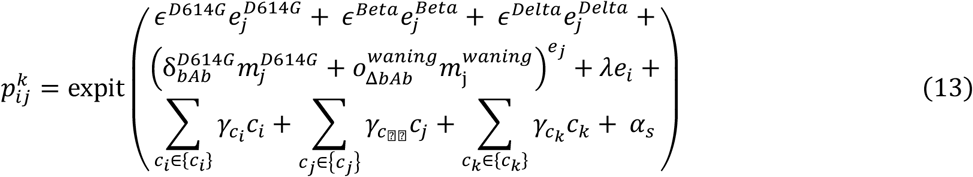

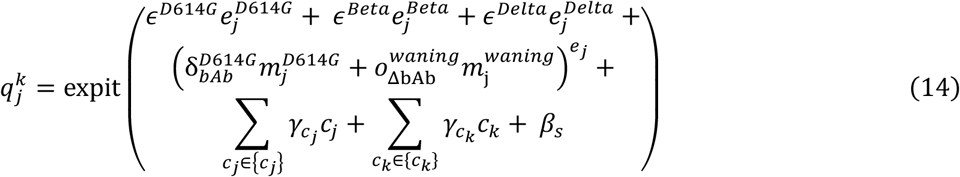

Here, 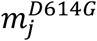 represents the D614G spike binding antibodies ELISA readout at blood draw 8 and 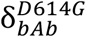 represents the effect size of 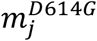 in mediating transmission probability 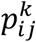 against the Omicron wave infection at blood draw 8. While 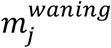 represents the drop from peak D614G spike binding antibodies readout prior to blood draw 8 (measured as the highest D614G spike binding Ab titer level among the first 8 blood draws) to that at blood draw 8 and 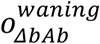 represents the effect size of 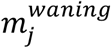 in mediating transmission probability 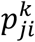 against the Omicron wave infection at blood draw 8. All other settings of Model 4 were kept the same as Model 2. The results of the Model 4 are presented in Extended Data Table 4.

We found that binding antibody levels at BD5 (BD8) correlate with protection against Delta (Omicron) wave infections: the risk of infection decreased by 74% (95% CI 41% – 88%) and 40% (95% CI 33% – 54%) per unit increase in OD value for the Delta and Omicron wave analyses, respectively. Conversely, the decline in bAbs from peak levels to BD5/BD8 (Δ*bAb*^*W*^) demonstrated no contribution to the overall protection, with risk reduction per 10-fold increase: - 2% (95%CI: -91% – 55%) for Delta wave infections and -2% (95%CI: -87% – 55%) for Omicron wave infections. These findings underscore the correspondence between waning of binding antibodies and a waning of protection. Furthermore, our estimations indicate that the proportion of protection conferred through D614G spike bAbs at BD5 is 35% (95%CI: 32% – 38%) against Delta wave infections, a figure comparable estimation based on anti-D614G nAbs (37%, 95%CI: 34% – 40%, Extended Data Table 4). Notably, D614G spike bAbs at BD8 accounted for 27% (95% CI: 25% – 29%) of protection against Omicron wave infection, representing a larger proportion compared to anti-BA.1 nAbs (11%, 95%CI: 9% – 12%, Extended Data Table 4).

#### Transmission chains imputation and parameters estimation based on an Expectation-maximization (EM) algorithm

Here we describe the process to fit the models described in Section 3 to the household infection data. The serologic data available for the Delta and Omicron only provides information on the total number of infections within the household between two blood draws collected before and after the SARS-CoV-2 wave. The data does not provide the details of the transmission chains within the household, the order of infections among infected individuals, nor the infection dates. To account for the uncertainties of the transmission tree structure within households given only the total number of infections, we enumerate and reconstruct all possible transmission chains among the infected individuals, where each infected individual may have been infected by members of their own household or the general community. Supplementary Fig. 1 illustrates all 16 possible configurations of transmission chains for a household with 3 infected individuals. We limited our analysis to households with no more than 6 infected individuals, as the possible configurations of transmission chains among 6 infected individuals already reaches 16,807. Enumeration of all possible transmission chain configurations would be computationally intractable for households with more than 6 infected individuals. Additionally, the probability of each possible transmission chain depends on the parameter estimates of the transmission model described in the previous session in Methods. To address the statistical uncertainties due to unresolved transmission chains (which would affect the statistical confidence of mediation analysis detailed in the prior section), we jointly fit the household transmission model and impute the topological structure of the transmission trees. We use an EM algorithm, as described below ^55^.

To resolve who infected whom within the household in a probabilistic manner, we considered an EM algorithm that iteratively estimates the transmission model parameters 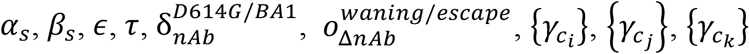 through maximizing the likelihood function *L* as described in Equation (1) in the previous section then updates the imputed probability of each transmission tree configuration within each household based on the fitted transmission model. The process is as follows:

1. Initial imputation of the household transmission trees with equal sampling probability for all configurations: For each household, we randomly sample one transmission tree with equal probability among all transmission tree configurations that are compatible with the number of infections. We iterate through all households so that each household has a simulated transmission tree. We then repeat the imputation 1000 times to obtain 1000 realizations of each household’s transmission tree.
2. Maximization step: We consider the waning parameter *τ* a hyper-parameter (nonlinear term in equations (3-6), cannot be estimated by logistic regression). For a fixed value of *τ*, for each of the 1000 realizations of the simulated household transmission chains, we estimate transmission model parameters 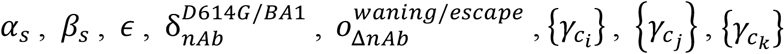 through maximizing the likelihood function *L* described in Equation (1). The maximization of the likelihood function is achieved through fitting a logistic regression of the infection/exposure outcomes for all participants using R package ***“brglm”*** (version 0.7.2). We then pool the estimates from the 1000 realizations using the ***“pool”*** function in the R package ***“mice”*** (version 3.16.0). The full likelihood of the combined Delta and Omicron waves fitting in this EM step *m* can be expressed as 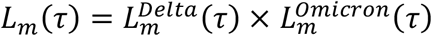
3. Expectation step: for a fixed value of hyper-parameter *τ*, based on the pooled estimates of the transmission model parameters 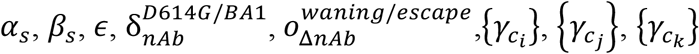 we calculate the likelihood all configurations of transmission chains within each household based on Equation (2). We use these configuration-specific likelihoods to resample transmission chains: For each household, we randomly sample one transmission tree among all transmission tree configurations with probability proportional to transmission tree likelihood prescribed in Equation (2), given the parameters estimated by the most recent maximization step. We iterate through all households so that each household is assigned one simulated transmission tree. We repeat the process 1000 times to obtain 1000 realizations of the household transmission trees.
4. For each of the fixed value of hyper-parameter *τ* over a plausible range (30 - 500 days), we iterate over the EM steps (2) and (3) until *L*_*m*_(*τ*) converge to the maximum value of the EM algorithm. We scan through the values of *τ* from 30 to 500 days at 10 days step. The EM algorithm convergence curve is shown in Supplementary Fig. 2 for each of the *τ* values. The EM algorithm converges at step 50, irrespective of the value of *τ*. The marginal likelihood of the model at *τ, L*(*τ*) is estimated by taking the average of *L*_*m*_(*τ*) for EM steps 50 through 100. Supplementary Fig. 3 shows the log of the likelihood *L*(*τ*) as a function of *τ*, based on a spline interpolation. The point estimate of *τ* is taken from the maximum of *l*og(*L*(*τ*)) while the 95% confidence interval is estimated by finding *τ* values with log-likelihood value at the maximum minus 1.92 (Supplementary Fig. 3).
5. We then take the best estimate of hyper-parameter *τ* and repeat the EM algorithm till convergence to estimate transmission model parameters 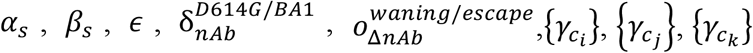 as show in Table 2. Same EM algorithm were applied to Model 2-4 for the sensitivity analysis as well.

The “treatment effect” by prior infection is estimated by simulating from the best-fit model. We first sample 1000 realizations of the imputed household transmission trees, with imputation probability proportional to the best estimates of transmission model using the EM algorithm and hyper-parameter *τ*. For each of the 1000 realizations, we focus on the subset of individuals who had one prior SARS-CoV-2 infection, denoted as *S*_*j*_ = {*j*| *e*_*j*_ = 1}). We use the fitted transmission model to predict the probability of infection (i.e. 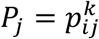 *or* 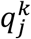, with) of these non-naïve subsets under three scenarios:

a. Scenario 1: the probability of infection estimated with predictors as reported in the data, denoted as 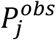.
b. Scenario 2: a counterfactual scenario where the probability of infection is estimated with predictor *e*_*j*_ = 0 (i.e. a counterfactual naïve individual) whereas all other covariates (confounders) are the same as observed, removing both direct and mediator effects. We denote the infection probability in this counterfactual scenario as 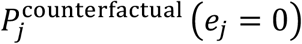.
c. Scenario 3: a counterfactual scenario where the probability of infection is estimated with predictor *e*_*j*_ = 1, but setting 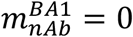 (or 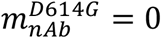), effectively removing the mediator effect of nAb on preventing transmission, but keeping the direct effect. We denote the infection probability in this counterfactual scenario as 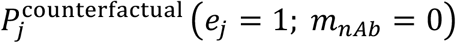.

We then calculate the total protection conferred by prior infection as the population average of 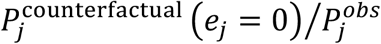, based on bootstrap resampling with replacement (maintaining the same number of observations) of each of the 1000 realizations of the household transmission chains. Point estimates and 95% confidence intervals are based on the median and 95% quantiles of 1000 realizations’ estimates.

Similarly, we calculate the proportion of protection mediated by nAbs as the population average of 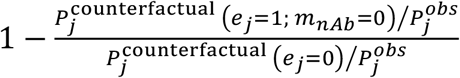. We use the same bootstrapping approach as for total protection.

## Data availability

Aggregate data to reproduce the figures are available at Zenodo (DOI: <u>10.5281/zenodo.11375487</u>). Individual-level data cannot be publicly shared because of ethical restrictions and the potential for identifying included individuals. Accessing individual participant data and a data dictionary defining each field in the dataset would require provision of protocol and ethics approval for the proposed use. To request individual participant data access, please submit a proposal to C.C <u>.</u> who will respond within 1 month of request. Upon approval, data can be made available through a data sharing agreement.

## Code availability

Code to reproduce the figures, using python version 3.8.11 and scipy version 1.7.1 is available at Zenodo (DOI: <u>10.5281/zenodo.11375487</u>).

## Extended Data Figures

**Extended Data Figure 1:**
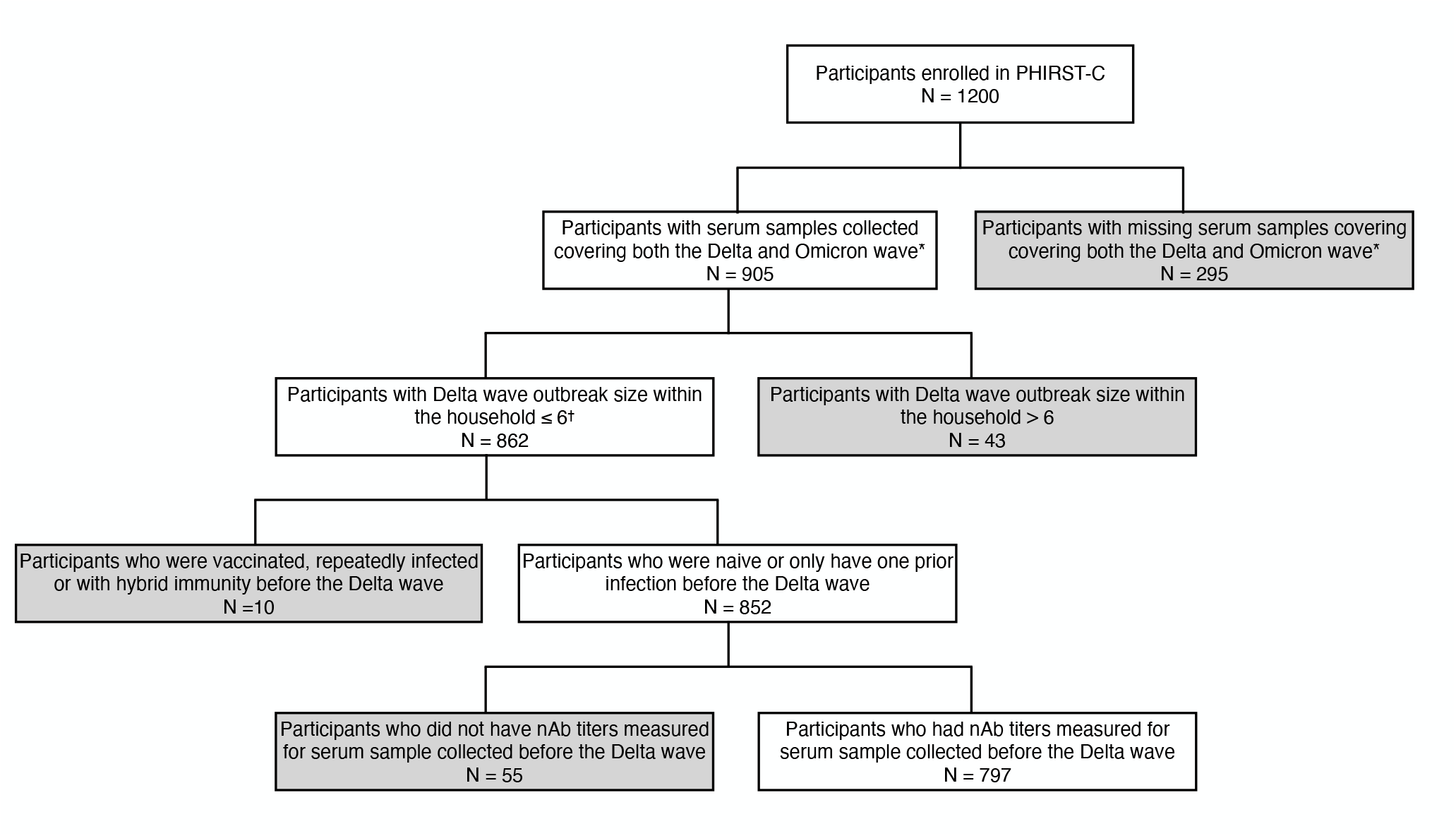
Flowchart of participants included in the Delta-wave subgroup analysis. Grey boxes represent participants excluded from the Delta-wave subgroup analysis. *Based on a previously published study ^30^. ^†^Household with more than 6 infected individuals would be computationally intractable to track all possible transmission chain configurations.

**Extended Data Figure 2:**
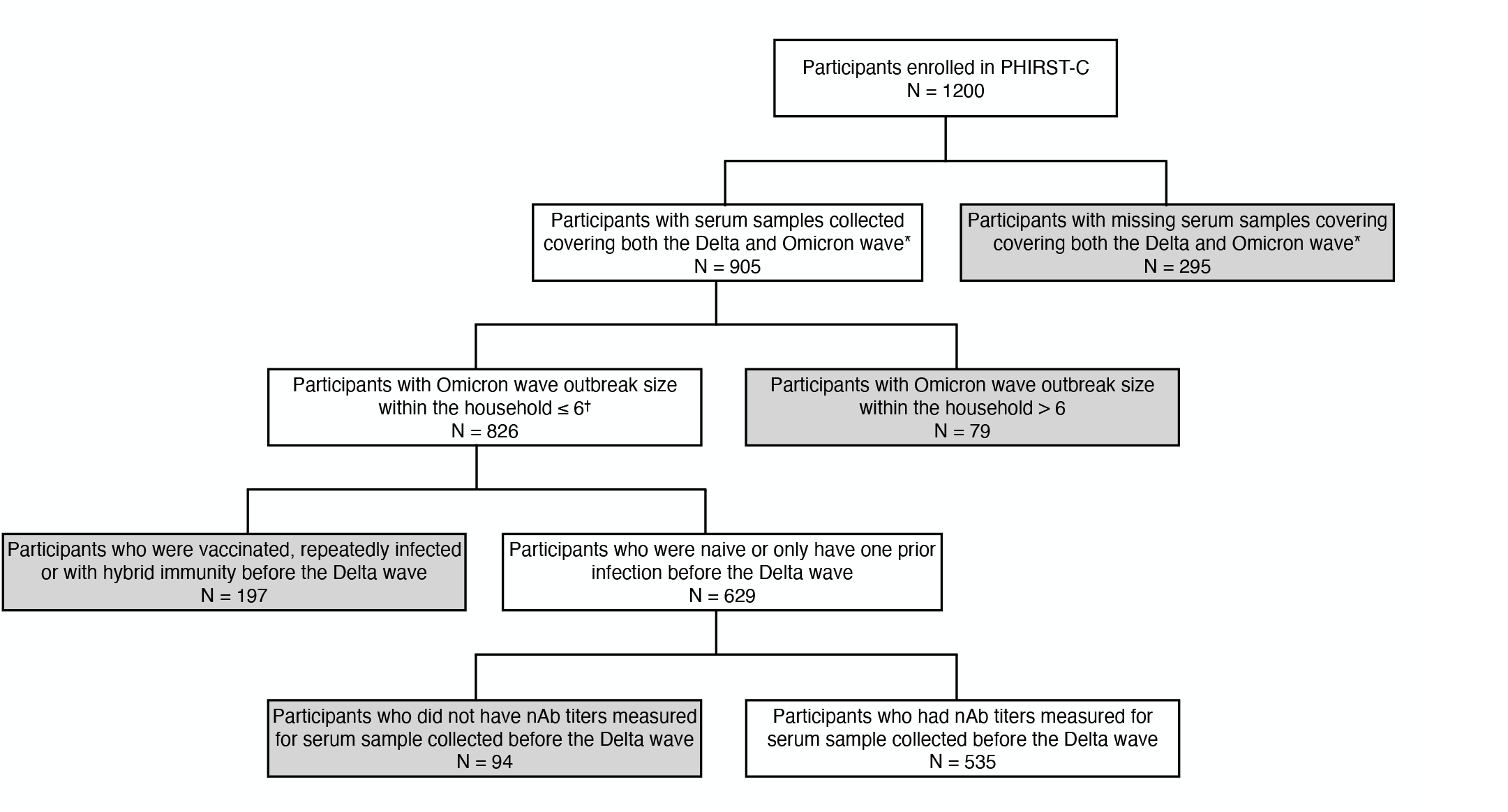
Flowchart of participants included in the Omicron-wave subgroup analysis. Grey boxes represent participants excluded from the Omicron-wave subgroup analysis. *Based on a previously published study ^30^. ^†^Household with more than 6 infected individuals would be computationally intractable to track all possible transmission chain configurations.

**Extended Data Figure 3:**
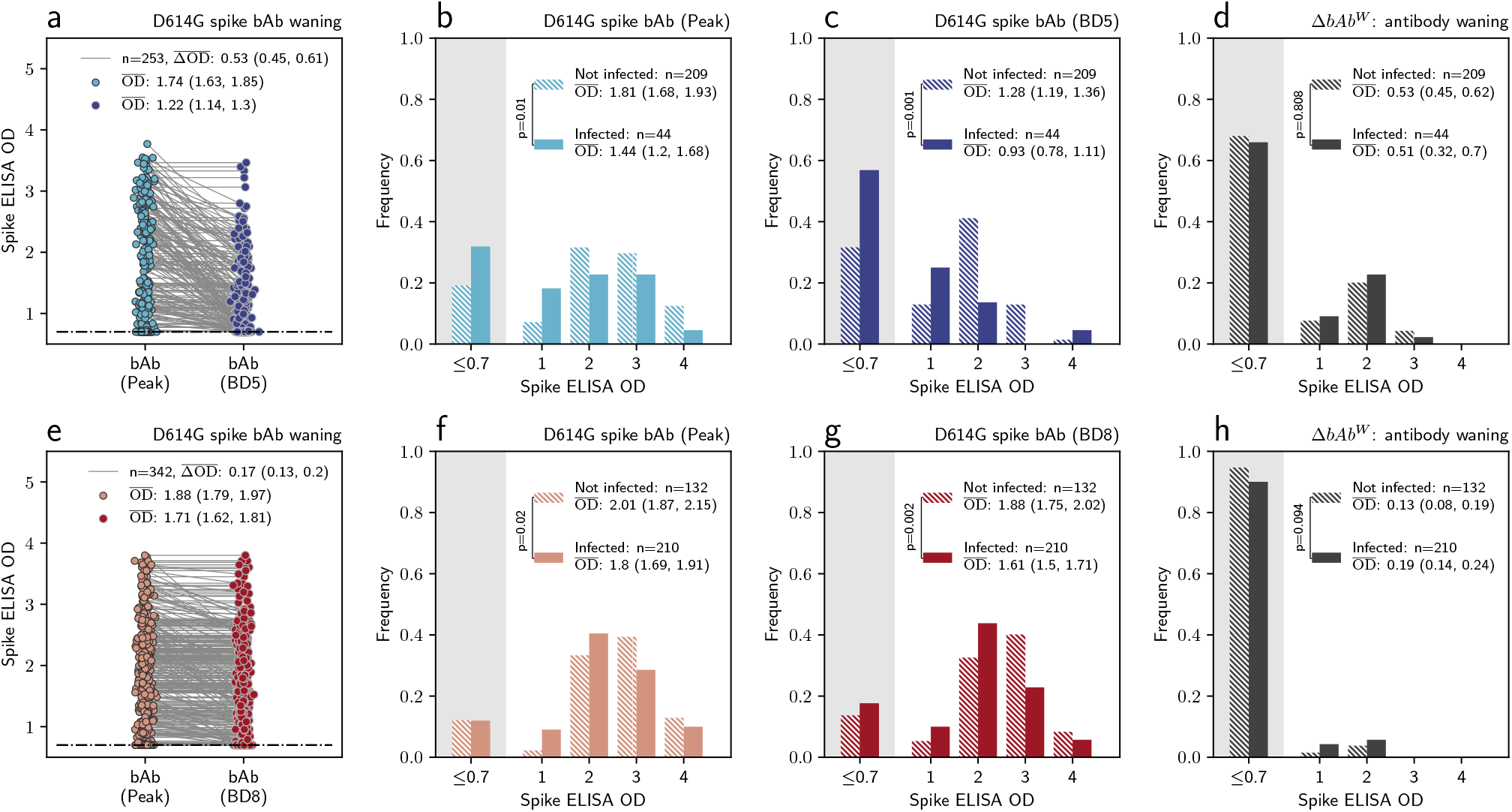
D614G spike binding antibody (bAb) level for the Delta wave and the Omicron wave analysis. **a**, for Delta wave subgroup, the distribution of the peak bAb level to BD5 (light blue dots) and the D614G spike bAb level at BD5 (dark blue dots), among individuals who had one prior SARS-CoV-2 infection before blood draw 5. Each dot represents one individual, with two measurements of the same individual connected through a gray line. OD: absorbance at 450 nm, measured in optical density 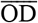: the average of OD; 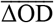: the average drop of OD. **b**, for Delta wave subgroup, the distribution of the peak D614G spike bAb up to BD5, stratified by individuals who were infected during the Delta wave (solid bar) vs those who were not infected (dashed bar). Independent samples t-test (two-sided) is used to determine the statistical significance (anti reported on the legend) of difference between the 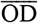 of the two groups. **c**, same as **b** but for D614G spike bAb level at BD5. **d**, same as **b** but for Δ*bAb*^*W*^. **e**, for Omicron wave subgroup, the distribution of the peak bAb level to BD8 (light red dots) and the D614G spike bAb level at BD8 (dark red dots), among individuals who had one prior SARS-CoV-2 infection before BD8. Each dot represents one individual, with two measurements of the same individual connected through a gray line. **f**, for the Omicron wave subgroup, the distribution of the D614G spike bAb level at BD8, stratified by individuals who were infected during the Omicron wave (solid bar) vs those who were not infected (dashed bar). Independent samples t-test (two-sided) is used to determine the statistical significance (p-value reported on the legend) of difference between the 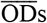 of the two groups. **g**, same as **f** but for D614G spike bAb level at BD8. **d**, same as **f** but for Δ*bAb*^*W*^.

## Extended Data Tables

**Extended Data Table 1:** Positivity rate of different serologic assays by the variant type of prior exposure for the Delta and Omicron wave subgroup.

| Delta wave subgroup |  |  |  |
| --- | --- | --- | --- |
| Seropositivity | Prior D614G infection | Prior Beta infection | Prior Delta exposure |
| Anti-nucleocapsid assay were positive in at least one of the first 5 blood draws | 109/113 (97%) | 133/140 (95%) | — |
| Anti-nucleocapsid assay were positive at BD5 | 104/113 (92%) | 129/140 (92%) | — |
| Anti-D614G nAb assay were positive for peak nAb response. | 87/113 (77%) | 60/140 (43%) |  |
| Anti-D614G nAb were positive for nAb response at BD5 | 81/113 (72%) | 59/140 (42%) | — |
| Omicron wave subgroup |  |  |  |
| Seropositivity | Prior D614G exposure | Prior Beta exposure | Prior Delta exposure |
| Anti-nucleocapsid assay were positive in at least one of the first 8 blood draws | 60/61 (98%) | 116/120 (97%) | 160/161 (99%) |
| Anti-nucleocapsid assay were positive at BD8 | 58/61 (95%) | 108/120 (90%) | 159/161 (99%) |
| Anti-D614G nAb were positive for nAb response at BD8 | 57/61 (93%) | 71/120 (59%) | 140/161 (87%) |
| Anti-BA.1 nAb were positive for nAb response at BD8 | 29/61 (48%) | 36/120 (30%) | 50/161 (31%) |

**Extended Data Table 2:** Mediation analysis for nAbs as CoPs against serologically ascertained Delta and Omicron wave infections, with a variant-specific model for direct effect. Average and 95% CIs are provided for each of the model parameters. Δ*nAb*^*W*^: the quantity of anti-D614G nAbs waned from peak level to that at BD5. Δ*nAb*^*E*^: the quantity of antibodies that can neutralize D614G but fail to neutralize Omicron BA.1 at BD8 due to Omicron’s immune escape.

| Wave |  |  | Delta | Omicron |
| --- | --- | --- | --- | --- |
| Protection against reinfection | Direct effect<br>(Protection absent of nAbs) | Prior D614G exposure<br>(odds ratio, absent of waning) | 0.76 (0.36, 1.61) | 1.23 (0.63, 2.38) |
|  |  | Prior Beta exposure<br>(odds ratio, absent of waning) | 0.47 (0.30, 0.76) | 0.78 (0.50, 1.21) |
|  |  | Prior Delta exposure<br>(odds ratio, absent of waning) | — | 0.47 (0.29, 0.76) |
|  | Mediators effect<br>(Protection from nAbs) | Anti-D614G nAb<br>(odds ratio, per 10-fold increase) | 0.59 (0.43, 0.83) | — |
| | | $\Delta nAb^W$<br>(odds ratio, per 10-fold increase) | 1.00 (0.72, 1.39) | — |
|  |  | Anti-BA.1 nAb<br>(odds ratio, per 10-fold increase) | — | 0.73 (0.56, 0.95) |
| | | $\Delta nAb^E$<br>(odds ratio, per 10-fold increase) | — | 0.94 (0.76, 1.15) |
|  | Total protection<br>(relative risk compared to naïve individuals) |  | 0.40 (0.38, 0.42) | 0.70 (0.68, 0.72) |
|  | Proportion of protection mediated by nAbs |  | 37% (34%, 40%) | 11% (9%, 12%) |
|  | Protection against onward transmission<br>(Odds ratio compared to naïve individuals) |  | 0.20 (0.05, 0.72) | 1.11 (0.62, 2.00) |

**Extended Data Table 3:**
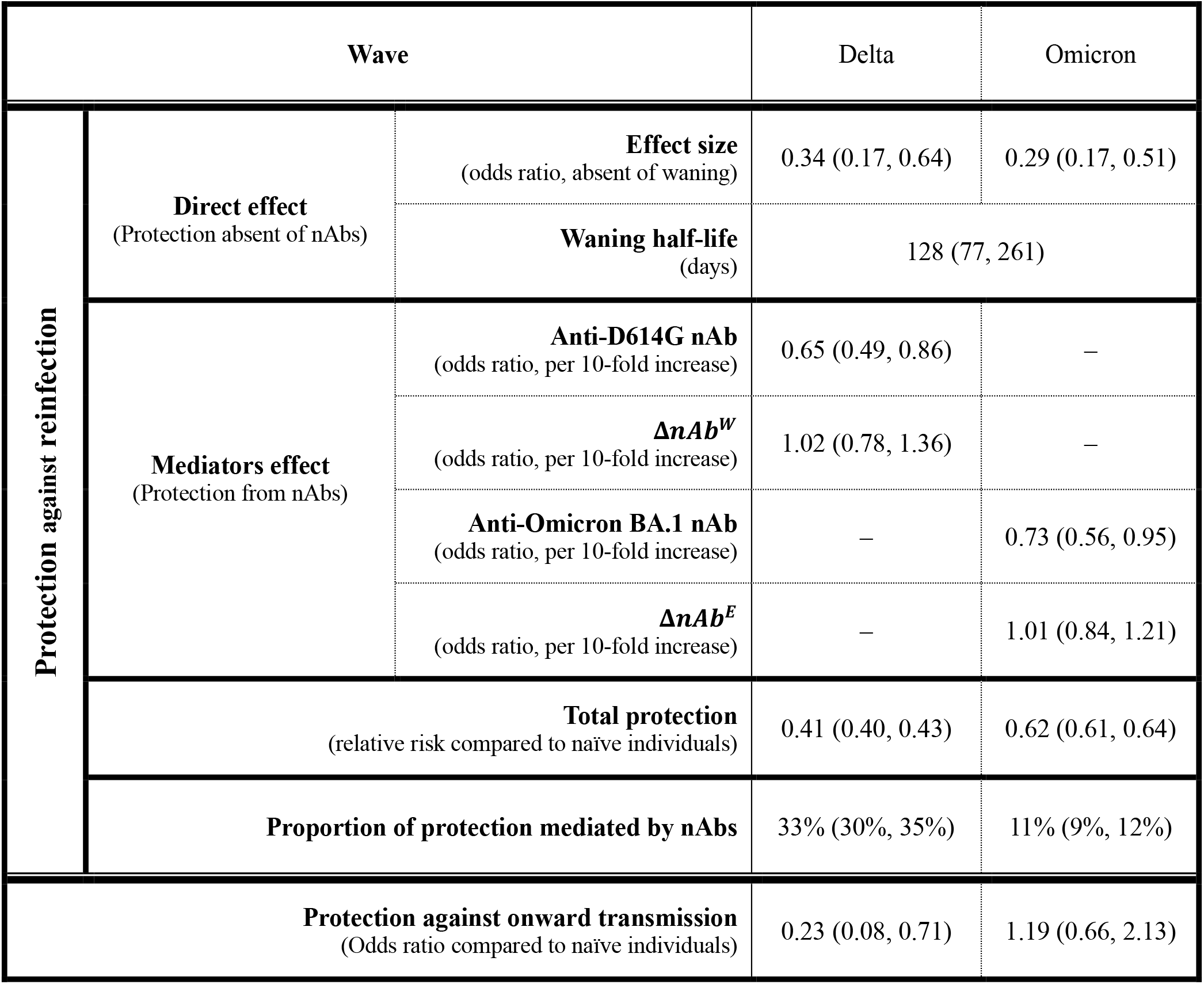
Mediation analysis for nAbs as CoPs against Delta (ascertained by both serology and PCR) and Omicron wave infections, with a waning model for direct effect. Average and 95% CIs are provided for each of the model parameters. Δ*nAb*^*W*^: the quantity of anti-D614G nAbs waned from peak level to that at BD5. Δ*nAb*^*E*^: the quantity of antibodies that can neutralize D614G but fail to neutralize Omicron BA.1 at BD8 due to Omicron’s immune escape.

**Extended Data Table 4:** Mediation analysis for D614G spike binding antibody as CoPs against serologically ascertained Delta and Omicron wave infections, with a variant-specific model for direct effect. Average and 95% CIs are provided for each of the model parameters. Δ*bAb*^*W*^: the quantity of D614G spike binding antibodies waned from peak level to that at BD5 for Delta (at BD8 for Omicron).

| Protection against reinfection |  |  | Delta<br>(serology) | Omicron<br>(serology) |
| --- | --- | --- | --- | --- |
|  | Direct effect<br>(Protection absent of nAbs) | Prior D614G exposure<br>(odds ratio, absent of waning) | 0.60 (0.24, 1.48) | 1.38 (0.67, 2.84) |
|  |  | Prior Beta exposure<br>(odds ratio, absent of waning) | 0.51 (0.32, 0.83) | 0.91 (0.58, 1.45) |
|  |  | Prior Delta exposure<br>(odds ratio, absent of waning) | — | 0.61 (0.38, 0.97) |
|  | Mediators effect<br>(Protection from nAbs) | D614G binding Ab<br>(odds ratio, per 10-unit increase) | 0.26 (0.12, 0.59) | 0.60 (0.46, 0.77) |
| | | $\Delta bAb^W$<br>(odds ratio, per 10-unit increase) | 1.02 (0.55, 1.91) | 1.02 (0.55, 1.87) |
|  | Total protection<br>(relative risk compared to naïve individuals) |  | 0.40 (0.38, 0.42) | 0.65 (0.63, 0.67) |
|  | Proportion of protection mediated by spike binding Ab |  | 35% (32%, 38%) | 27% (25%, 29%) |
| Protection against onward transmission<br>(Odds ratio compared to naïve individuals) |  |  | 0.22 (0.06, 0.74) | 1.18 (0.65, 2.13) |

## Supplementary Information

**Supplementary Fig. 1:**
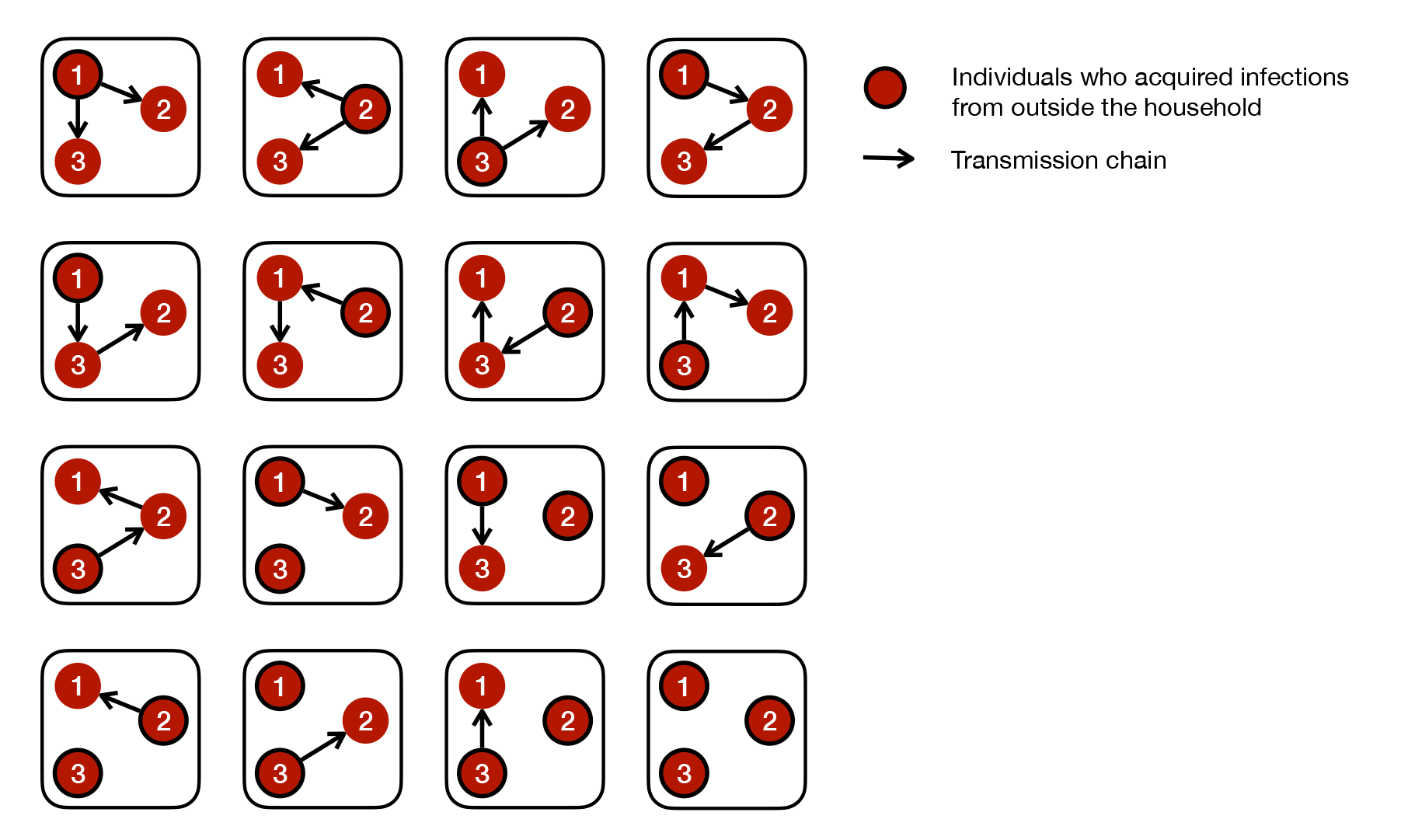
Visualization of all 16 possible transmission chains within a household of 3 infected individuals.

**Supplementary Fig. 2:**
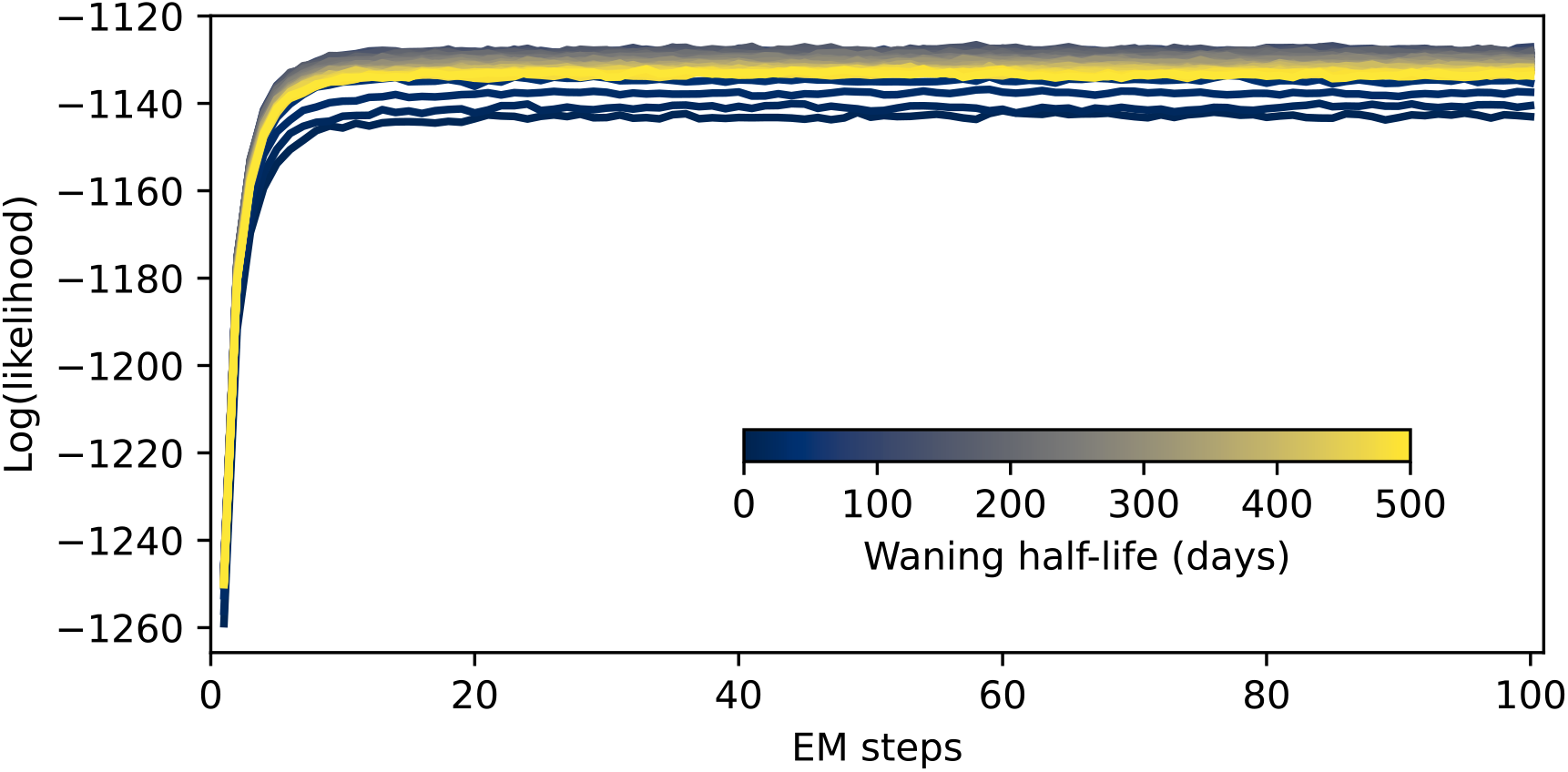
The log-likelihood of the transmission model fit, as a function of the EM steps.

**Supplementary Fig. 3:**
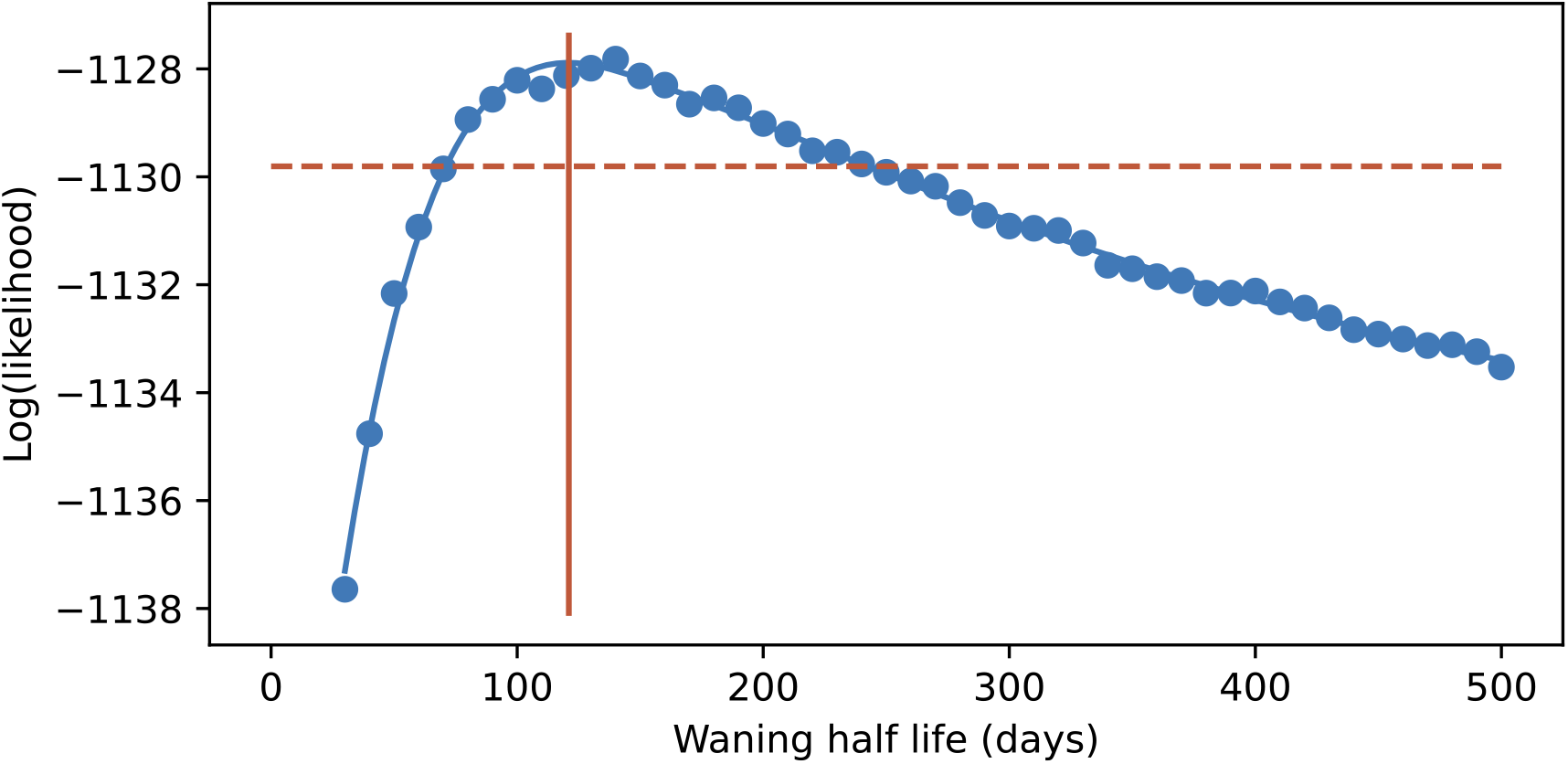
The profile log-likelihood of the transmission model over hyper-parameter of waning half-life.

Solid vertical line indicates the waning half-life corresponding to maximum of the profile likelihood. Dashed horizontal line represent 1.92 below the maximum profile likelihood.

